# Cortical surface area is an independent predictor of incident dementia

**DOI:** 10.64898/2026.11.13.26367749

**Authors:** Haley E. Wiskoski, Summan Zahra, Scott R. French, Juan C. Arias, Xana Waughman, Loi Do, Cris Escareno, Paola Vidana, Raza Mushtaq, Kevin Johnson, Maria Altbach, Eric M. Reiman, Fanny M. Elahi, Francesca Vitali, Edward J. Bedrick, Theodore P. Trouard, Gene E. Alexander, Craig C. Weinkauf, the Alzheimer’s Disease Neuroimaging Initiative

## Abstract

Cortical surface area (cSA) is associated with intelligence and cognition, yet structural MRI studies of neurodegeneration in Alzheimer’s disease and related dementias (ADRD) have emphasized quantification of cortical thickness (cTH) and volume, with very limited evaluation of cSA. We examined whether cSA of AD-relevant brain regions provide diagnostic, prognostic, and biological information beyond cTH and volume across 5,430 participants (3,124 cognitively unimpaired; 1,634 mild cognitive impairment [MCI]; 672 ADRD) from three aging and dementia cohorts: the National Alzheimer’s Coordinating Center, Alzheimer’s Disease Neuroimaging Initiative, and Carotids and Minds study. In cross-sectional analyses, cSA and cTH showed weak correlation, and lower cSA was independently associated with greater odds of MCI and ADRD after adjustment for cTH and covariates. Participants with both low cSA and low cTH had greater risk of MCI and ADRD compared to low cSA, cTH, or volume alone. In longitudinal models, baseline cSA predicted incident ADRD independent from cTH, and combined cSA/cTH models outperformed cSA, cTH, or volume alone for predicting incident ADRD over a mean follow-up of 4.3 years. Complementing these findings, proteomics analyses revealed distinct signatures associated with cSA compared to cTH. Overall, these findings support cSA as a clinically informative and biologically distinct dimension of AD-related cortical vulnerability.

## Introduction

Alzheimer’s disease (AD) is characterized by neurodegeneration and a progressive decline in cognitive function, with structural magnetic resonance imaging (MRI) playing a central role in quantifying disease-relevant brain atrophy.^1^ MRI-derived cortical thickness (cTH) and volume are widely used markers of neurodegeneration in AD and related dementias (ADRD),^2^ providing non-invasive measures of cortical atrophy. Cortical thinning within AD-vulnerable regions commonly referred to as the AD-Signature^3^ has been shown to distinguish prodromal AD from cognitively unimpaired controls, identify individuals with AD-like biomarker profiles, and predict risk of ADRD-related cognitive decline.^4–6^ These findings have established cTH as a robust MRI marker of ADRD-related neurodegeneration and have reinforced a broader tendency to conceptualize cortical atrophy primarily through thinning.

Cortical surface area (cSA) has received comparatively less attention in aging and dementia research, despite cortical volume being a composite measure of both cTH and cSA, two metrics reflecting distinct dimensions of cortical morphology. cSA has been linked to cognition and intelligence across the lifespan^7–9^ and has been shown in some populations to have stronger influence on cortical volume than cTH.^10,11^ Developmental models link cortical surface expansion to progenitor proliferation and tangential expansion of cortical columns, whereas cTH reflects radial growth and layer formation within the cortical sheet.^12,13^ Twin and genetic studies of adolescent participants further demonstrate that cSA and cTH are highly heritable but genetically distinct.^10,14,15^ Together, these findings indicate that volume combines separable sources of biological variation, and evaluating these dimensions separately may provide a more precise framework for interpreting cortical morphometry in brain aging than volume or cTH alone. In the context of ADRD, we hypothesized that cSA may reflect a clinically relevant dimension of cortical vulnerability that is not captured by thinning.

Although previous studies suggest that surface-based morphometrics may capture information not fully represented by cTH,^16–19^ the evidence remains limited and inconsistent. A previous cross-sectional study showed that cSA and cTH differ across cognitively unimpaired controls (CU), mild cognitive impairment (MCI), and AD groups but was limited by sample size and did not test whether cSA adds independent diagnostic or prognostic value beyond cTH or volume.^16^ Longitudinal studies reported distinct cSA, cTH, and volume atrophy patterns in early AD^17^ and across diagnostic groups^18,19^ but were limited to short follow-up intervals, small sample size, whole-brain regional assessment, and did not determine whether cSA predicts clinical diagnosis or incident ADRD independent of other established morphometric measures. Furthermore, it has not been established whether cSA is independent of cTH, whether combined cSA/cTH phenotypes stratify ADRD risk, or whether cSA adds diagnostic or prognostic value beyond cTH or volume. Beyond this, whether cSA and cTH are associated with distinct molecular/cellular pathways remains understudied in neurodegeneration. Such evidence could help determine whether these measures reflect distinct biological pathways in aging and neurodegeneration.

To address these gaps, we examined cSA, cTH, and volume of AD-relevant brain regions using 5,430 participants drawn from three cohorts:^20–24^ the National Alzheimer’s Coordinating Center (NACC), Alzheimer’s Disease Neuroimaging Initiative (ADNI), and University of Arizona Carotids and Minds study (CAM). The AD-Signature and Default Mode Network (DMN) regions were selected as they have been related to early cortical changes associated with ADRD.^3,25,26^ Through cross-sectional and longitudinal evaluation, we assessed whether cSA provides clinically relevant prognostic and diagnostic information independent of cTH, and whether their combined evaluation provides clinically meaningful information beyond cTH or volume alone. We further assessed whether cSA and cTH were associated with overlapping or distinct proteomic signatures in ADNI, providing molecular and cellular context for their associations with ADRD-related outcomes. This multi-cohort study aims to determine whether cSA represents a distinct and clinically informative dimension of cortical vulnerability across the ADRD spectrum.

## Materials and methods

### Participants

Participants were drawn from three cohorts: the National Alzheimer’s Coordinating Center (NACC), Alzheimer’s Disease Neuroimaging Initiative (ADNI) and University of Arizona Carotids and Minds study (CAM). The ADNI (adni.loni.usc.edu) was launched in 2003 as a public-private partnership, led by Principal Investigator Michael W. Weiner, MD. The original goal of ADNI was to test whether serial magnetic resonance imaging (MRI), positron emission tomography (PET), other biological markers, and clinical and neuropsychological assessment can be combined to measure the progression of mild cognitive impairment (MCI) and early Alzheimer’s disease (AD). The National Alzheimer’s Coordinating Center (NACC) was established in 1999 by the National Institute on Aging (NIA) and serves as the centralized data repository for the ADRCs. NACC and ADNI were used as large multisite aging and AD cohorts to evaluate cross-sectional diagnostic associations and progression to incident AD.^20,21^ CAM was included as a single-centre, vascular-enriched cohort to evaluate demographic and vascular disease associations with morphometry in participants enriched for extracranial carotid atherosclerotic disease (ECAD) and vascular risk.^22–24^

For NACC and ADNI, participants >50 years with at least one available structural MRI were included (Supplemental Fig. 1). Exclusions included neurological or psychiatric disease/disorder other than MCI or ADRD, serious or unstable medical illness, and missing demographic or diagnostic data. In NACC, MRI visits were required to be within 180 days of clinical diagnosis. Baseline cognitive status was categorized as cognitively unimpaired (CU), mild cognitive impairment (MCI), or ADRD according to cohort-specific diagnostic procedures including NACC Uniform Data Set procedures^27,28^ and ADNI diagnostic criteria.^21^ In NACC, impaired-not-MCI and MCI groups were combined, and the ADRD group included both AD-dementia and vascular dementia. Longitudinal analyses were restricted to ADNI participants who were CU or MCI at baseline with at least one follow-up MRI assessment (Supplemental Fig. 2). In ADNI, status of hyperlipidaemia, smoking history, and diabetes were extracted from medical history text fields. In-depth ADNI protocols can be found at https://adni.loni.usc.edu/help-faqs/adni-documentation/.

The CAM study recruited participants aged 50-85 through cardiology and vascular surgery clinics across Tucson, AZ for an ongoing study of vascular disease association with dementia risk.^22–24^ Inclusion criteria include ECAD and/or at least two vascular risk factors (hypertension, hyperlipidaemia, history of smoking, or diabetes). ECAD was defined as having a percent carotid artery stenosis greater than or equal to 50%, determined using the North America Symptomatic Carotid Endarterectomy Trial criteria and time-of-flight carotid MRI as described in a previous study.^22^ Study exclusion criteria included a clinical diagnosis of ADRD or other neurological disorders, stroke within six months, major medical illness, or major depression. Participants with large cortical infarcts were further excluded from this study (Supplemental Fig. 3). CAM does not include clinical/cognitive staging. All participants in the current study provided written informed consent in accordance with institutional review board guidelines.

### MRI Data Acquisition & Processing

Structural T1-weighted MRI data were obtained according to cohort-specific protocols. ADNI MRI was acquired across ADNI phases using standardized protocols at 1.5T and 3T;^29^ both original and pre-processed images (including gradient distortion and intensity inhomogeneity correction) were utilized.^30^ Both 1.5T and 3T MRI from ADNI were included to increase sample size. NACC MRI was acquired at participating Alzheimer’s Disease Research Centers;^31^ imaging used from the NACC cohort was acquired at 3T and not pre-processed. CAM imaging was acquired on a single 3T scanner (Magnetom Skyra, Siemens Healthineers) using a previously described protocol.^32^

Cortical reconstruction and morphometric estimation were performed using *FreeSurfer v7* using the standard pipeline with a single T1-weighted image as input.*^33^* NACC morphometrics were derived from T1-weighted images by NACC using *FreeSurfer* v7.2, ADNI morphometrics by the Banner Alzheimer’s Institute (Phoenix, AZ) research group using *FreeSurfer* v7, and CAM morphometrics locally using *FreeSurfer* v7.3. Primary measures included cSA, cTH and volume. cSA and volume were computed as bilateral region sums, and cTH as the bilateral mean.

The primary region of interest was an AD-Signature cortical composite, defined using *FreeSurfer* Desikan-Killiany atlas regions implicated in AD neurodegeneration (Figure 1A): entorhinal cortex, inferior temporal cortex, middle temporal cortex, temporal pole, inferior parietal cortex, superior parietal cortex, praecuneus and posterior cingulate cortex.^3,34^ Temporal pole was unavailable in NACC and omitted from that cohort’s composite. Secondary analyses using a Default Mode Network (DMN) composite were also performed given established vulnerability in ADRD.^25,26^

**Figure 1.**
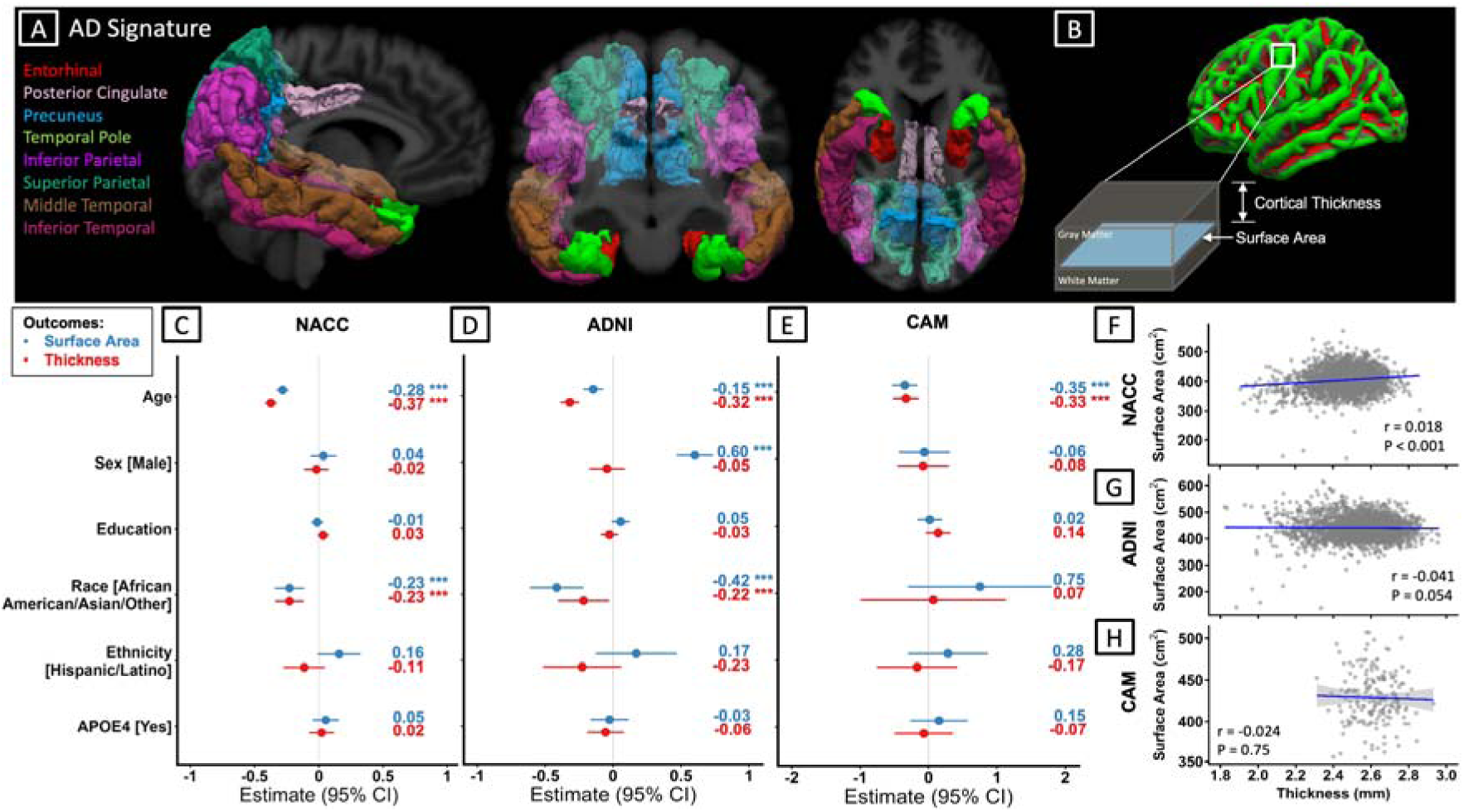
AD-Signature Cortical Regions and Associations with Demographic Characteristics Across Cohorts. **A)** AD-Signature cortical regions included in the composite measure. **B)** Schematic illustrating cSA and cTH as components of cortical morphology. **C-E)** Forrest plots illustrating multiple linear regression standardized β estimates and 95% CIs for associations of demographic and genetic factors among CU participants in NACC, ADNI, and CAM cohorts. In ADNI, the model was additionally adjusted for magnetic field strength and image preprocessing status. **F-H)** Scatterplots showing correlations between AD-Signature cSA and cTH across the full diagnostic spectrum in NACC, ADNI, and CAM. Error bars represent 95% CIs. \**P*<0.05; \*\**P*<0.01; \*\*\**P*<0.001. Abbreviations: AD, Alzheimer’s disease; ADNI, Alzheimer’s Disease Neuroimaging Initiative; APOE4, apolipoprotein E ε4 allele; CAM, Carotids and Minds; CI, confidence interval; cSA, cortical surface area; cTH, cortical thickness; CU, cognitively unimpaired; NACC, National Alzheimer’s Coordinating Center.

cSA and volume were adjusted for total intracranial volume (TIV) within each cohort using residualization with CU participants serving as reference. cTH was not TIV-adjusted. Outlier detection was performed per cohort and metric: Whole brain cSA or cTH values exceeding ±3 standard deviations from the cohort mean were flagged, and regional morphometric values were reviewed to identify biologically plausible variation, data entry errors, or processing artifacts. Participants with implausible or missing morphometric values were excluded.

### CSF Proteomics

We obtained SomaScan 7k panel (v4.1)^35^ data for 7008 proteins which were simultaneously assayed in CSF from 694 ADNI participants with available cSA and cTH measurements at their baseline MRI. The SomaScan assay utilizes modified single-stranded DNA aptamers to bind protein targets with high sensitivity and specificity, which are quantified by DNA microarray.^36,37^ Proteins are measured in relative fluorescent units (RFU). Initial data normalization was performed by Somalogic,^35^ while assay and quality-control procedures were performed by the Cruchaga lab at WashU as previously described.^35^

### Statistical Analysis

All analyses were conducted independently in NACC, ADNI and CAM. Correlations between cSA and cTH were assessed within each cohort using Spearman correlation. Demographic associations with cSA and cTH were evaluated using multivariable linear regression models. Association of cSA and cTH with MCI and ADRD were evaluated using cross-sectional multivariable logistic regression comparing CU vs. MCI and ADRD as outcomes. All models were adjusted for age, sex, years of education, race, ethnicity, and *apolipoprotein E* (*APOE*) ε4 carrier status; ADNI models additionally included magnetic field strength and image pre-processing status. *β* estimates for all regression models were standardized. In models estimating risk associated with lower morphometrics, measures were multiplied by -1 so that odds/hazard ratios greater than 1 reflected higher odds/hazard per 1-standard deviation lower morphometric value. Secondary models were performed using the DMN as a regional composite, and ADNI sensitivity analyses were performed restricting the sample to participants whose morphometrics were derived from 3T, un-processed imaging.

To evaluate cross-sectional cSA/cTH phenotypes, NACC and ADNI cohort-specific lower-quartile thresholds were used to classify participants as normal cSA/normal cTH, normal cSA/low cTH, low cSA/normal cTH, and low cSA/low cTH, with “low” defining values below the threshold, and “normal” defining values above the lower-quartile threshold. This was performed instead of using the highest quartile to define “normal” in order to maximize sample size. Similar was done to delineate high/low volume. Multivariable logistic regressions were used to evaluate the association of morphometric phenotypes with MCI and ADRD compared to CU using normal cSA/normal cTH as the reference factor. Additional models were assessed exchanging the reference factor to assess differences between phenotype groups. To assess cross-sectional discrimination across phenotypes, receiver operating characteristic (ROC) analyses were performed for CU vs. ADRD.

Associations of baseline cSA and cTH to incident ADRD were assessed using Cox proportional hazards models among ADNI participants who were CU or MCI at baseline. The proportional hazards assumption was assessed using scaled Schoenfeld residuals for each model. Time-to-event was defined as time from baseline MRI to first observed ADRD diagnosis. Participants who did not progress to ADRD were censored at the last available MRI follow-up visit date. All Cox models included age, sex, education, race, ethnicity, *APOE* ε4 carrier status, magnetic field strength and imaging preprocessing status as covariates.

Nested Cox models were used to assess whether cSA provided incremental prognostic information beyond cTH and whether decomposed morphometry outperformed cortical volume. Likelihood ratio tests compared nested models including covariates alone (age, sex, education, race, ethnicity, *APOE* ε4 carriage, magnet strength, image preprocessing status), covariates plus cSA, covariates plus cTH, covariates plus volume, covariates plus cSA and cTH, and covariates plus cSA, cTH, and volume. Model performance was summarized using Akaike information criterion (AIC) and Harrel C-index.

Optimized cSA, cTH, and volume cutoffs were derived within the longitudinal ADNI cohort using ROC curve analyses among participants who were CU/MCI at baseline. For each measure, ROC curves were used to evaluate the discrimination of incident ADRD. Optimal thresholds were defined using Youden’s J statistic. Participants were classified as having normal/low cSA, cTH, and volume, based on the threshold maximizing Youden’s J statistic. Binary cSA/cTH classifications were combined into four phenotypes: normal cSA/normal cTH, normal cSA/low cTH, low cSA/normal cTH, and low cSA/low cTH which were used in subsequent survival analyses. Kaplan-Meier curves were generated for normal/low cSA, normal/low cTH, normal/low volume, and combined cSA/cTH phenotype groups, with differences assessed using log-rank tests. Statistical significance was defined as *P*<.05 in all analyses. Testing was performed in *R* version 4.4.3 using the following packages: *survival, survminer.* ^38,39^

ADNI CSF proteomics data were analysed using multiple linear regression models adjusted for age, sex, race, ethnicity, and MRI magnet strength to identify differentially expressed proteins (DEPs) associated with cSA and cTH. DEPs were visualized using volcano plots and protein-protein interaction (PPI) networks. PPI networks were constructed using STRING (12.0). PPIs had a minimum confidence of >0.4 (cSA) and >0.7 (cTH). Overrepresentation analysis (ORA) was performed with DEPs as input and 7008 SomaScan proteins as background to identify pathways associated with cSA and cTH using *ShinyGO* (0.85.1).^40^ The Reactome and Gene Ontology (GO) gene sets were queried.^37^ Pathways containing 10-500 proteins were considered significantly enriched. All *P* values were corrected for multiple comparisons using Benjamini Hochberg False Discovery Method (FDR, *P* <0.05).

## Results

### Participants

2,993 NACC, 2,255 ADNI, and 182 CAM participants were included in the study (Supplemental Table 1). Participants were older adults with mean ages of 71±8 years in NACC, 73±7 in ADNI, and 72±7 in CAM. Female sex was 63% in NACC, and 47% in both ADNI and CAM. *APOE* ε4 allele was present in 38% of NACC, 46% of ADNI, and 27% of CAM with available genotype data. Vascular risk factors (hypertension, diabetes, hyperlipidaemia and smoking) had greater prevalence in CAM, consistent with its vascular-enriched study design. NACC included a larger proportion of CU participants (CU=2,094; MCI=605; ADRD=294), while ADNI had a larger proportion of MCI and ADRD (CU=848; MCI=1,029; ADRD=378) (Supplemental Table 1). The CAM study did not involve clinical/cognitive staging.

### Associations of cSA and cTH with Demographic and Genetic Factors

In CU participants, AD-Signature cSA and cTH (Figure 1A-B) showed associations with demographic and genetic factors across cohorts (Figure 1C-E). Older age was consistently associated with significantly lower cSA and cTH while education, ethnicity and *APOE* ε4 did not show any significant associations. Non-White racial groups were associated with lower cSA and cTH in both NACC and ADNI. Male sex was associated with greater cSA in ADNI. 1.5T magnetic field strength and pre-processed imaging (including gradient distortion and intensity inhomogeneity correction) were associated with lower cSA in ADNI (Supplemental Fig. 4). Field strength and image preprocessing status were consistent across NACC and CAM and was thus not assessed. Full demographic and neuroimaging covariate associations are shown in Supplemental Figure 1. Similar demographic findings were observed in the DMN-associated brain regions and for the ADNI sensitivity analysis restricted to 3T, unprocessed imaging (Supplemental Fig. 5-6).

The correlations between cSA and cTH were near zero in the full cohorts of NACC (*r*=0.018, *P*<0.001), ADNI (*r*=-0.041, *P*=0.054) and CAM (*r*=-0.024, *P*=0.75) (Figure 1F-H). Similar was observed in the DMN (Supplemental Fig. 7). Among CU, MCI and ADRD participants, *APOE* ε4 allele presence was associated with lower cTH and volume in NACC and ADNI, and lower cSA only in ADNI (Supplemental Fig. 8).

Analyses in CU participants showed no consistent association between common vascular risk factors and cSA across NACC, ADNI, and CAM (Supplemental Fig. 9). Hypertension was weakly associated with lower cTH in NACC and ADNI (*β*=-0.12 and *β*=-0.14, respectively; both *P*<0.05). In CAM, carotid artery disease was associated with lower cSA (*β*=-0.52; *P*<0.01) but not cTH; carotid disease was not evaluated in NACC and ADNI. None of the vascular risk factors showed significant independent association with cSA or cTH in CAM.

### Associations of cSA and cTH with Cognitive Status

Mean cSA and cTH differed significantly across cognitive status (CU, MCI, ADRD) in NACC and ADNI (all *P*<0.001; Figure 2A-B). In multivariable logistic regression models, lower cSA was associated with higher odds ratio (OR) of MCI and ADRD independent of cTH and other covariates (age, sex, education years, race, ethnicity, *APOE* ε4 presence, and magnetic field strength and image preprocessing status in ADNI) in both NACC (MCI: OR, 1.53; ADRD: OR, 2.49; *P*<0.001) and ADNI (MCI: OR,1.48; ADRD: OR, 2.14; *P*<0.001) (Figure 2C-D). Lower cTH was also independently associated with higher odds of MCI and ADRD in NACC (MCI: OR, 1.67; ADRD: OR, 6.18; *P*<0.001) and ADNI (MCI: OR, 2.28; ADRD: OR, 9.25; *P*<0.001). Full model results are provided in Supplemental Fig. 10. Similar associations were observed for DMN morphometry, and associations remained significant in the ADNI sensitivity analysis restricted to 3T unprocessed imaging (Supplemental Figure 11-13).

**Figure 2.**
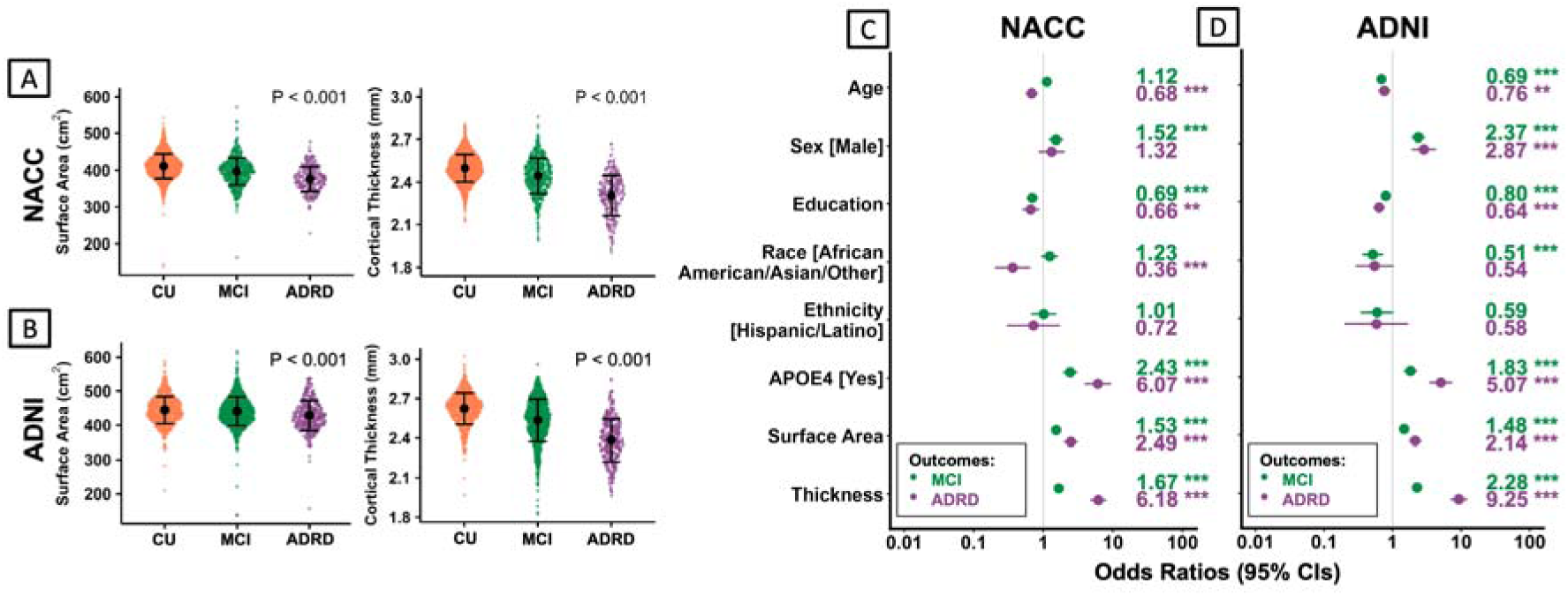
AD-Signature Surface Area and Thickness Differ Across Diagnostic Groups with Independent Associations with MCI and ADRD. A-B) Distributions of AD-Signature cSA and cTH across diagnostic groups (CU, orange; MCI, green; ADRD, purple) in NACC and ADNI. Points represent individual participants; mean and standard deviations are shown. **C-D)** Multivariable logistic regression models showing odds ratios and 95% CIs for association of cSA and cTH to MCI (green) and ADRD (purple) relative to CU participants in NACC and ADNI. Models were adjusted for age, sex, education, race, ethnicity, and *APOE* ε4 status. The ADNI model was additionally adjusted for magnetic field strength and image preprocessing status. cSA and cTH were standardized and directionally coded so that odds ratios greater than one indicate higher odds with lower morphometric values. \**P*<0.05; \*\**P*<0.01; \*\*\**P*<0.001. Abbreviations: ADNI, Alzheimer’s Disease Neuroimaging Initiative; ADRD; Alzheimer’s disease and related dementias; APOE4, apolipoprotein E ε4 allele; CI, confidence interval; cSA, cortical surface area; cTH, cortical thickness; CU, cognitively unimpaired; MCI, mild cognitive impairment; NACC, National Alzheimer’s Coordinating Center.

### Associations of Morphometric Phenotypes with Cognitive Status – Normal vs. Low cSA and cTH

Cross-sectional combined cSA/cTH phenotypes were defined by lower-quartile thresholds in NACC and ADNI (Supplemental Fig. 14). In both cohorts, ADRD participants made up a majority of the low cSA/low cTH phenotype (53% for both). Phenotype distributions within the DMN brain regions and ADNI sensitivity sub-cohort also followed this pattern (Supplemental Fig. 15-16). In adjusted logistic regression models, low cSA/low cTH, low cSA/normal cTH and normal cSA/low cTH were associated with higher odds of MCI and ADRD relative to the normal cSA/normal cTH group (Figure 3A-B). Low cSA/low cTH showed significantly greater odds than either isolated low cSA or low cTH, with the strongest odds of clinical impairment across all phenotypes in NACC (MCI: OR, 4.47; ADRD: OR, 86.58; *P*<0.001) and ADNI (MCI: OR, 10.28; ADRD: OR, 83.13; *P*<0.001). Low volume also showed significantly greater odds of MCI and ADRD relative to normal volume in NACC and ADNI, but odds ratios were numerically less than the low cSA/low cTH phenotype (Figure 3A-B). In area under curve (AUC) analyses, cSA/cTH phenotypes demonstrated strong discrimination of ADRD from CU in NACC and ADNI, where low cSA/low cTH showed the greatest classification performance and exceeded low volume alone (Figure 3C-D). Full phenotype model results, DMN analyses, and ADNI sensitivity analyses are provided in Supplemental Figures 17-21.

**Figure 3.**
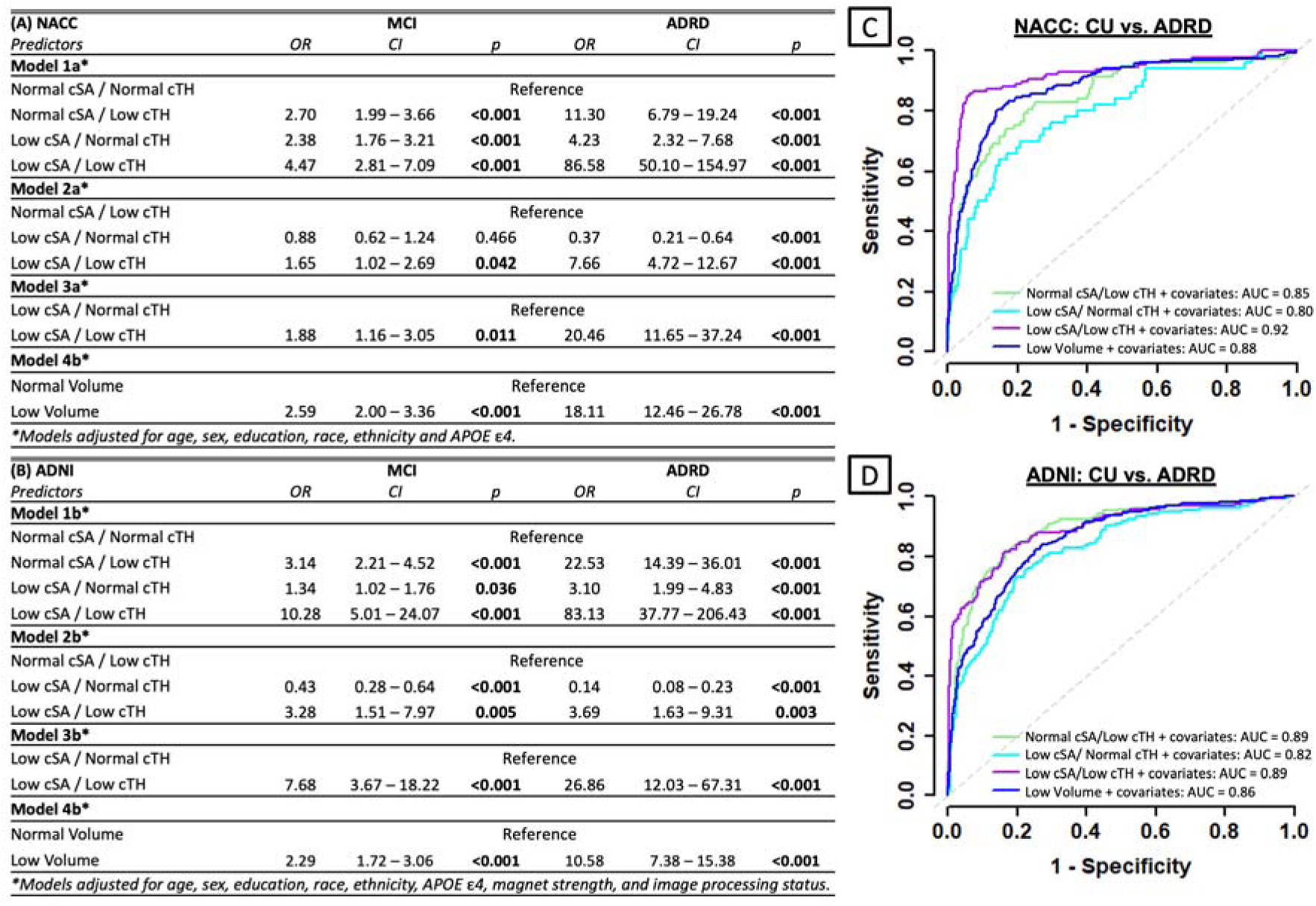
Combined Low cSA/Low cTH Phenotype Shows Strongest Association with MCI and ADRD. Multivariable logistic regression models showing odds ratios and 95% CIs for MCI and ADRD relative to CU participants in **(A)** NACC and **(B)** ADNI. Participants were classified into normal or low AD-Signature cSA and cTH groups based on lower-quartile cut points. NACC models were adjusted for age, sex, education, race, ethnicity, and *APOE* ε4 status. ADNI models were additionally adjusted for magnetic field strength and image pre-processing status. ROC curves showing classification of CU vs ADRD in **(C)** NACC and **(D)** ADNI. Models included covariates and AUC values are shown for each phenotype. Abbreviations: AD, Alzheimer’s disease; ADNI, Alzheimer’s Disease Neuroimaging Initiative; ADRD; Alzheimer’s disease and related dementias; APOE4, apolipoprotein E ε4 allele; AUC, area under the curve; CI, confidence interval; cSA, cortical surface area; cTH, cortical thickness; CU, cognitively unimpaired; MCI, mild cognitive impairment; NACC, National Alzheimer’s Coordinating Center; OR, odds ratio; ROC, receiver operating characteristic.

### Association of cSA and cTH with Longitudinal ADRD Progression

Longitudinal analyses included 1,558 ADNI participants who were CU or MCI at baseline (mean age: 73±7 years, 46% female, 42% *APOE* ε4 carriers, 42% CU); mean follow-up span of MRI observations from baseline was 4.3±3.1 years with a maximum follow-up span of 15.8 years (Supplemental Table 2). 336 participants had a conversion to ADRD over their follow-up span. NACC was not evaluated longitudinally as sample size was smaller for CU/MCI participants having at least one follow-up MRI visit (*n*=754) and mean follow-up span was lower (1.7±0.8 years, max=4.2 years) with only 44 ADRD conversion events. In ADNI, lower baseline cSA and cTH were independently and significantly associated with greater risk of progression to incident ADRD (Supplemental Fig. 22). In a secondary analysis restricted to participants who were CU at baseline, lower baseline cSA and cTH were also independently and significantly associated with greater risk of progression to incident MCI (Supplemental Fig. 22).

In nested Cox proportional hazards models predicting incident ADRD, addition of cSA significantly improved the model fit beyond demographic, genetic, and imaging covariates, including age, sex, education, race, ethnicity, *APOE* ε4 allele presence, field strength, and image pre-processing status (likelihood ratio test [LRT] χ²=25.41; *P*<0.001; Supplemental Fig. 23). cSA remained informative after accounting for cTH in the model: adding cSA to the cTH-adjusted model significantly improved model fit (LRT χ²=18.46; *P*<0.001). Compared to the volume model, the cTH and cSA model had a lower Akaike Information Criterion (AIC) (ΔAIC=59.94) and greater Harrel C-index (0.81 vs. 0.78; Supplemental Fig. 23).

Models including cortical morphology measures of both cSA and cTH outperformed models based on cortical volume as the sole morphometric predictor. Adding cSA and cTH to a volume-adjusted model significantly improved fit (LRT χ²=64.24; *P*<0.001), whereas adding volume to the cSA and cTH model did not result in substantial improvement (LRT χ²=2.30; *P*=0.13, Supplemental Fig. 23). Consistent with this, discrimination was essentially unchanged after adding volume to the cSA + cTH model (ΔAIC=0.30; Supplemental Fig. 23).

Youden’s J-derived cSA, cTH and volume cutoffs for phenotype delineation and associated cohort distributions are provided in Supplemental Fig. 24-25. Kaplan-Meier analyses of baseline CU/MCI participants demonstrated significant differences in ADRD-free survival by cSA, cTH and combined cSA/cTH phenotype (all log-rank *P*<0.001; Figure 4A-C). Participants with low cSA or low cTH had lower ADRD-free survival compared with those in the normal groups (Figure 4A-B). When cSA and cTH were modelled jointly, the low cSA/low cTH phenotype showed the steepest decline in ADRD-free survival, whereas the normal cSA/normal cTH group showed the highest ADRD-free survival probability (Figure 4C). The low cSA/low cTH and low volume phenotypes were both associated with significantly poorer ADRD-free survival (both log-rank *P*<0.001), with low cSA/low cTH showing a steeper decline in survival probability (Figure 4D, Supplemental Figure 26).

**Figure 4.**
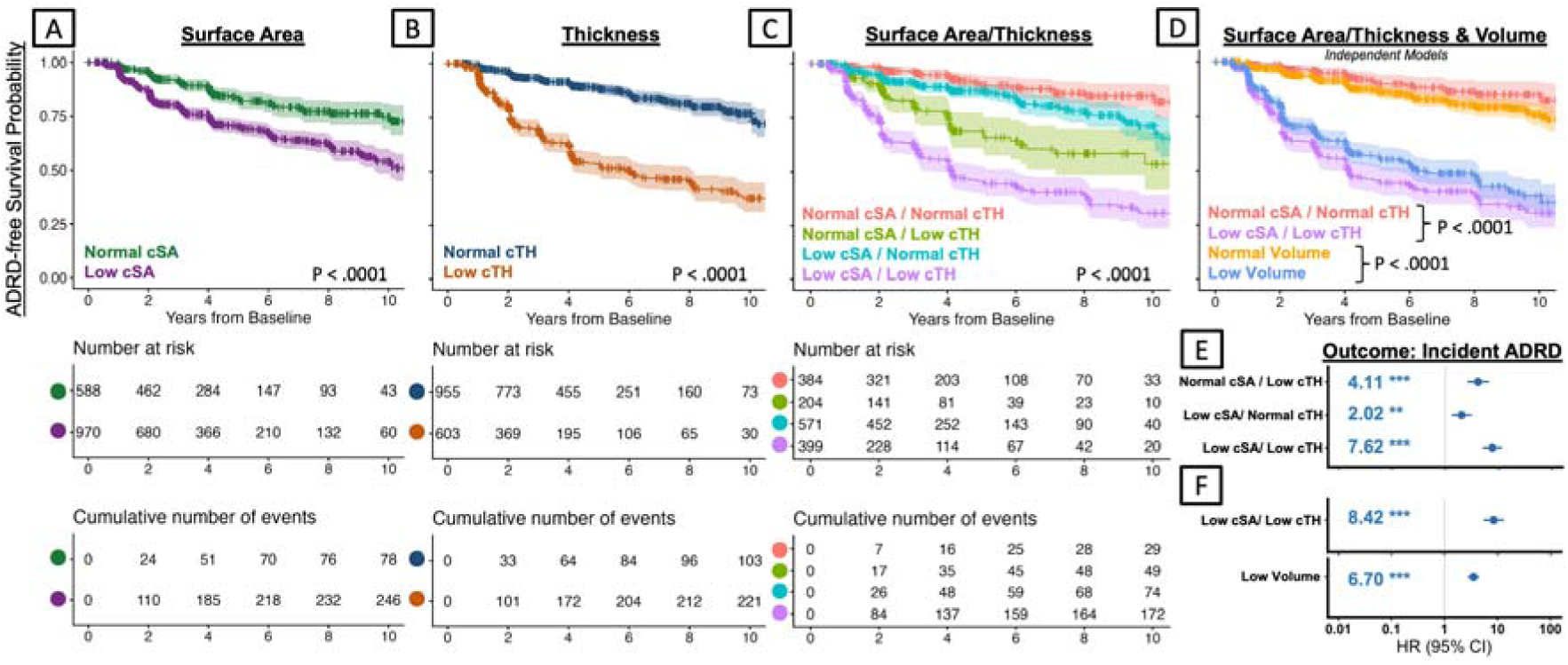
Baseline AD-Signature Morphometry and Risk of Incident ADRD in ADNI. A-D) Kaplan-Meier curves showing ADRD-free survival by baseline AD-Signature cSA, cTH, volume, and combined cSA/cTH phenotype. Shaded areas indicate 95% CIs; tables show numbers at risk and cumulative incident ADRD events over the follow-up time. Years from baseline are clipped at 10 years for visualization purposes. **E-F)** Cox proportional hazards models for incident ADRD including cSA/cTH phenotypes or volume group. Models were adjusted for age, sex, education, race, ethnicity, *APOE* ε4 status, magnet strength, image preprocessing status. Normal cSA/normal cTH or normal volume was the reference category where applicable. Hazard ratios greater than one indicate higher risk of incident ADRD. \**P*<0.05; \*\**P*<0.01; \*\*\**P*<0.001. Abbreviations: AD, Alzheimer’s disease; ADNI, Alzheimer’s Disease Neuroimaging Initiative; ADRD; Alzheimer’s disease and related dementias; CI, confidence interval; cSA, cortical surface area; cTH, cortical thickness; HR, hazard ratio.

In multivariable Cox proportional hazard models, baseline cSA/cTH phenotype was independently associated with greater risk of incident ADRD after adjustment for demographic, genetic, and imaging covariates (Figure 4E). Compared to the normal cSA/normal cTH group, normal cSA/low cTH was associated with a 4.11-fold higher hazard of incident ADRD, low cSA/normal cTH with a 2.02-fold higher hazard, and low cSA/low cTH with a 7.62-fold higher hazard (all *P*<.01). Direct comparison of low cSA/low cTH, excluding intermediate phenotypes from the model, showed a significantly higher hazard of incident ADRD than normal cSA/normal cTH (hazard ratio [HR], 8.42; *P*<0.001), where low volume was also associated with increased ADRD risk, although with a numerically smaller effect size (HR, 6.70; *P*<0.001) (Figure 4F). Full models in AD-Signature and DMN are shown in Supplemental Figures 27-28.

### CSF Proteomic Signatures of cSA and cTH

CSF proteomics was available for 694 ADNI participants (mean age 74±7, 43% female, 49% *APOE* ε4 carriers, 20% ADRD); the mean interval between CSF collection and baseline MRI was 89±281 days (Supplemental Table 3). Serum proteomics were only available in a smaller subset and were thus not assessed in this study. 67 Differentially Expressed Proteins (DEPs) were identified for cSA while 169 DEPs were identified for cTH, only 12 of which were shared (Supplemental Fig. 29). Gene Ontology Biological Processes (GO-BP) enrichment analysis revealed distinct biological pathways associated with cSA and cTH (Figure 5, Supplemental Fig. 29). Down-regulated proteins associated with semaphorin receptor signalling, ephrin receptor signalling, MET regulation, and Wnt signalling were associated with lower cSA (Figure 5, Supplemental Fig. 30, Supplemental Data), while these pathways were not significant for cTH. Down-regulated proteins associated with calcineurin signalling, RAB geranylgeranylation, 14-3-3 adaptor protein–IGF pathway interactions and synaptic MAP/Erk/Akt pathway interactions were associated with reduced cTH (Figure 5). Functional pathways of synapse assembly and axonogenesis were shared between cSA and cTH (Figure 5). A full summary of proteins and pathways associated with cSA and cTH is provided in Supplemental Data. Unaltered protein-protein interaction networks are shown in Supplemental Figure 30.

**Figure 5:**
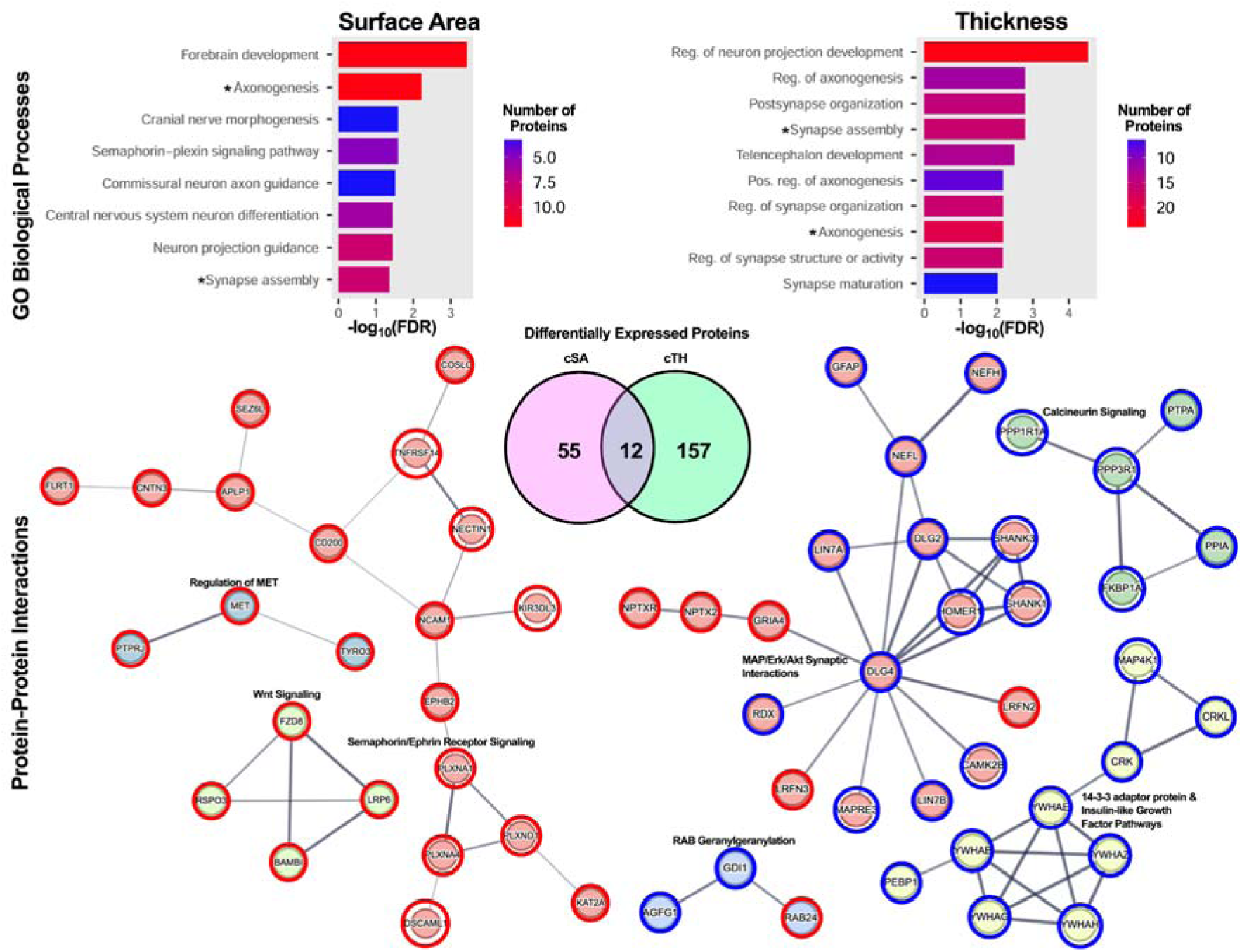
Cortical Surface Area and Thickness Have Unique Proteomic Signatures. Multiple linear regression models adjusted for age, sex, race, ethnicity, and MRI magnet strength were used to identify proteins associated with cSA and cTH in ADNI participants, which were then visualized using protein-protein interaction networks (PPIs) and used as inputs for pathway enrichment analysis with the GO Biological Processes database. The full SomaScan 7K menu was used as background for enrichment analysis. Proteins and pathways with FDR *P*<0.05 were deemed significant. PPIs had a minimum confidence of >0.4 (cSA) and >0.7 (cTH). Nodes are coloured according to clusters of related proteins and notable pathways are labelled in bold. Shared pathways are depicted with an asterisk. Red circles denote positive association with cSA or cTH, where blue circles denote negative association with cSA or cTH. Abbreviations: ADNI, Alzheimer’s Disease Neuroimaging Initiative; cSA, cortical surface area; cTH, cortical thickness; FDR, false discovery rate.

## Discussion

The majority of ADRD structural neuroimaging studies to date have focused on cTH and volume, reflecting strong historical evidence that ADRD is associated with cortical thinning and regional atrophy.^3,5,41–43^ Although cortical volume is a composite measure of cSA and cTH, few studies have evaluated cSA as an independent factor of cortical vulnerability. In this multi-cohort study across vascular-enriched and memory clinic cohorts, we demonstrated that cSA in the AD-Signature and DMN regions provide clinically relevant information not captured by cTH or volume alone. Lower cSA was independently associated with MCI and ADRD cross-sectionally and predicted incident ADRD over a mean follow-up of 4.3 years. Moreover, individuals with both low cSA and low cTH showed the greatest clinical vulnerability i.e., the highest risk for MCI and ADRD, supporting the value of decomposing cortical volume into its component dimensions. Together, the results of this study suggest that cortical vulnerability in ADRD is better represented when evaluating cSA in addition to cTH.

Multiple aspects of our data support conceptualization and evaluation of cSA and cTH as highly independent morphometric features that may reflect distinct biological vulnerabilities. cSA and cTH were only weakly correlated in all three cohorts, and both metrics showed independent associations with MCI, ADRD, and incident ADRD. The proteomics analyses also support this concept through demonstration of distinct cellular and molecular pathways in cSA and cTH. Although cSA has not been as well studied in the aging and dementia fields, evaluating cSA as an independent feature of brain morphometry is in keeping with developmental biology studies that have shown cSA and cTH are highly heritable traits with distinct genetic pathways.^10,14,15^ Moreover, these findings are consistent with evolutionary and developmental studies that have shown greater cSA is associated with higher intelligence and cognitive ability in humans^7–9^ and with mammalian studies suggesting that evolutionary expansion of cSA supports greater cognitive capacity across species.^44,45^ Together, these findings underscore that cSA, like cTH, represents a fundamental and unique dimension of cortical architecture.

From a clinical perspective, combined cSA/cTH phenotyping may provide a practical framework for morphometric risk stratification. Cortical volume and cTH are commonly used for prognostication of dementia risk, however our data suggest that evaluating cSA and cTH together improves identification of individuals at greater risk for clinical progression. Participants with low cSA and low cTH had the poorest ADRD-free survival and the highest risk of incident ADRD than either morphometric dimension alone. One possible interpretation is that low cSA may reflect a more developmentally constrained or premorbid dimension of cortical vulnerability, where low cTH may more closely reflect active neurodegenerative tissue loss. Under this framework, the low cSA/low cTH phenotype may represent a subgroup with more extensive or multifactorial cortical damage, helping explain its stronger association with ADRD progression than either metric alone.

The added value of cSA/cTH phenotyping may reflect, in part, the limitations of cortical volume as a composite measure. Because volume reflects both dimensions, similar volumetric values can arise from different cSA/cTH profiles that may not carry the same biological or clinical meaning; collapsing these into a single aggregate volume measure may mask this separable morphometric information. Consistent with this interpretation, models incorporating both cSA and cTH demonstrated better prognostic performance than models using cTH or volume alone and adding volume to a cSA+cTH model did not significantly improve model fit. Mathematically, the combined measure may also disproportionally reflect whichever dimension accounts for more variance, and it has been suggested that more interindividual variation in cortical volume is attributed to cSA rather than cTH.^10,11^ Furthermore, a composite measure makes it difficult to distinguish additive versus combined interactive effects that may hinder causal inference if cSA and cTH have different associations with the outcome of interest. As such, decomposing volume into cSA and cTH may therefore improve not only prognostic performance, but also biological interpretability.

Risk factor analyses further suggest that cSA and cTH may not be equally sensitive to all sources of dementia risk. Across cohorts, older age was consistently associated with lower cSA and cTH, consistent with prior work showing age-related reductions in both cortical dimensions.^46,47^ *APOE* ε4 allele presence was not significantly associated with either morphometric dimension in CU participants, but full cohort (with MCI and ADRD participants) analyses showed an association with lower cTH, consistent with previous studies linking *APOE* ε4 to cortical thinning.^48,49^ However, we found no consistent association between *APOE* ε4 allele presence and cSA. General vascular risk factors showed inconsistent associations with cortical morphometry, suggesting that broad vascular risk burden may not map uniformly onto cSA or cTH. Hypertension was associated with lower cTH in NACC and ADNI, in agreement with prior reports linking vascular risk to cortical thinning,^50–54^ but this was not observed in CAM. Notably, carotid disease in CAM participants was associated with significantly lower cSA, but not cTH, highlighting a potentially distinct pattern related to cerebrovascular disease. These distinctions are interesting because they indicate hypertension, *APOE* ε4 allele presence, age, and carotid stenosis may influence cortical structure through different mechanisms or timescales, and these differences highlight the benefit of assessing both cSA and cTH and the need for further research into cSA/cTH interactions. Because ADNI and NACC do not evaluate large vessel cerebrovascular disease, the extent to which carotid stenosis or other measures of vascular burden contribute to cortical morphometry in these cohorts remains unknown and will require further investigation.

The proteomics analyses provide added biological insight into the morphometric and clinical findings. Proteomics findings were broadly in agreement with published findings regarding pathways relevant in cTH deficits,^55^ the majority of which were not associated with cSA. Shared pathways significant in both cSA and cTH included axonogenesis and synapse assembly, pathways well-established in ADRD-related neurodegeneration.^56–59^ cSA had several distinct molecular/cellular pathway associations that may help explain why it contributed information beyond cTH and volume. One such pathway associated with decreased cSA is reduced ephrin/semaphorin signalling, which has established relevance in mutual repulsion of Cajal-Retzius (CR) cells, promoting final dispersion of CR cells over the cortical surface during neurodevelopment.^60^ Defects in migration of CR cells, controlled by Eph/Ephrin and Sema/Plexin pathways, can impair cortical aerealization.^61^ Relevant to ephrin/semaphorin signalling, preclinical studies have shown that amyloid beta oligomers can directly trigger degradation of EphB2 leading to reduced synaptic plasticity,^62^ while loss of PLXNA4/PLXND1 is associated with memory impairment^63^ and blood-brain barrier injury,^64^ linking reduced ephrin/semaphorin signalling to pathways of early neurodegeneration. We also found an association between decreased cSA and reduced Wnt signalling, an important pathway for cellular proliferation; loss of Wnt signalling has been associated with synaptic loss and amyloidogenic processing of amyloid precursor protein in preclinical studies of AD.^65^ These results further support our findings that cSA and cTH play independent roles in cortical vulnerability, reflecting distinct biological pathways that may differentially contribute to ADRD-related neurodegeneration.

Several methodological features relevant to prior gaps in the literature add to the rigor of this study. First, the use of a large sample drawn from three separate cohorts increases generalizability and reduces concern that the findings reflect cohort or acquisition/processing-specific biases. However, not every analysis could be performed with each dataset due to cohort-specific limitations in variable of interest availability. Second, cSA and cTH were examined within a priori AD-Signature and DMN brain region composites rather than through unconstrained regional testing across the whole cortex reducing multiple-comparison burden and grounding the analysis in regions with established ADRD vulnerability. Third, longitudinal analyses with up to 15-year follow-up provide evidence that baseline cSA relates to future clinical progression. Finally, CSF proteomic analyses offer biological context for the distinct associations seen in cSA and cTH.

This study also has several limitations. 1) The observational design precludes causal inference, and lower cSA/cTH may reflect preexisting vulnerability or other accumulated disease-related differences. 2) Although models were adjusted for key demographic, genetic, and imaging covariates, residual confounding of comorbidities and other cohort-specific characteristics remains possible. 3) Morphometric estimates rely on *FreeSurfer* segmentation accuracy, which may be dependent on participant age and post-processing protocols^32^. 4) Normal/low phenotypes are useful for visualization and risk stratification but are cohort-derived and should thus be interpreted as pragmatic summaries rather than fixed biological thresholds. 5) Our longitudinal analyses show that baseline cSA and cTH predict future clinical progression, but do not assess cSA or cTH over time. Future studies could determine whether cSA is primarily a vulnerability marker, dynamic disease marker, or both. 6) Analyses did not separately evaluate progression from CU to MCI and MCI to ADRD or stratify these transitions by evidence of amyloid pathology; therefore, incremental prognostic value of cSA beyond cortical volume, *APOE* ε4, and baseline cognition within biomarker-defined clinical subgroups remains uncertain. Future studies incorporating longitudinal morphometric measurements, stage- and biomarker-specific analyses, and independent replication are needed to address these limitations and further clarify the biological and prognostic significance of cSA.

In conclusion, these findings support cSA as a clinically relevant dimension of cortical morphology. Future studies should consider decomposing cortical volume into cSA and cTH to better characterize distinct and complementary patterns of cortical vulnerability. Incorporating cSA into ADRD neuroimaging frameworks may improve risk stratification, clarify heterogeneous pathways towards dementia, and provide a stronger morphometric bridge to studies of neurodegeneration.

## Supporting information

Supplemental CSF proteomics results.

## Data availability

The deidentified CAM data utilized in the analyses for this study will be made available to qualified researchers upon reasonable request to the corresponding author (CW) and receipt of a signed data access agreement form. Data from the ADNI and NACC are not owned or distributed by the study authors. ADNI data can be requested through the LONI Image and Data Archive (https://adni.loni.usc.edu/data-samples/adni-data/#AccessData), subject to approval and adherence to the ADNI Data Use Agreement. NACC data can be requested through the NACC data request process (naccdata.org/data-request-process), subject to approval and adherence to the NACC Data Use Agreement.

## Acknowledgements

The authors thank Yi Su, PhD, and Vivek Devadas of Banner Alzheimer’s Institute (Phoenix, Arizona) for providing the FreeSurfer-derived cortical morphometric data from the Alzheimer’s Disease Neuroimaging Initiative (ADNI) used in this study.

