## Supplemental Figures and Tables. for "Cortical surface area is an independent predictor of incident dementia"

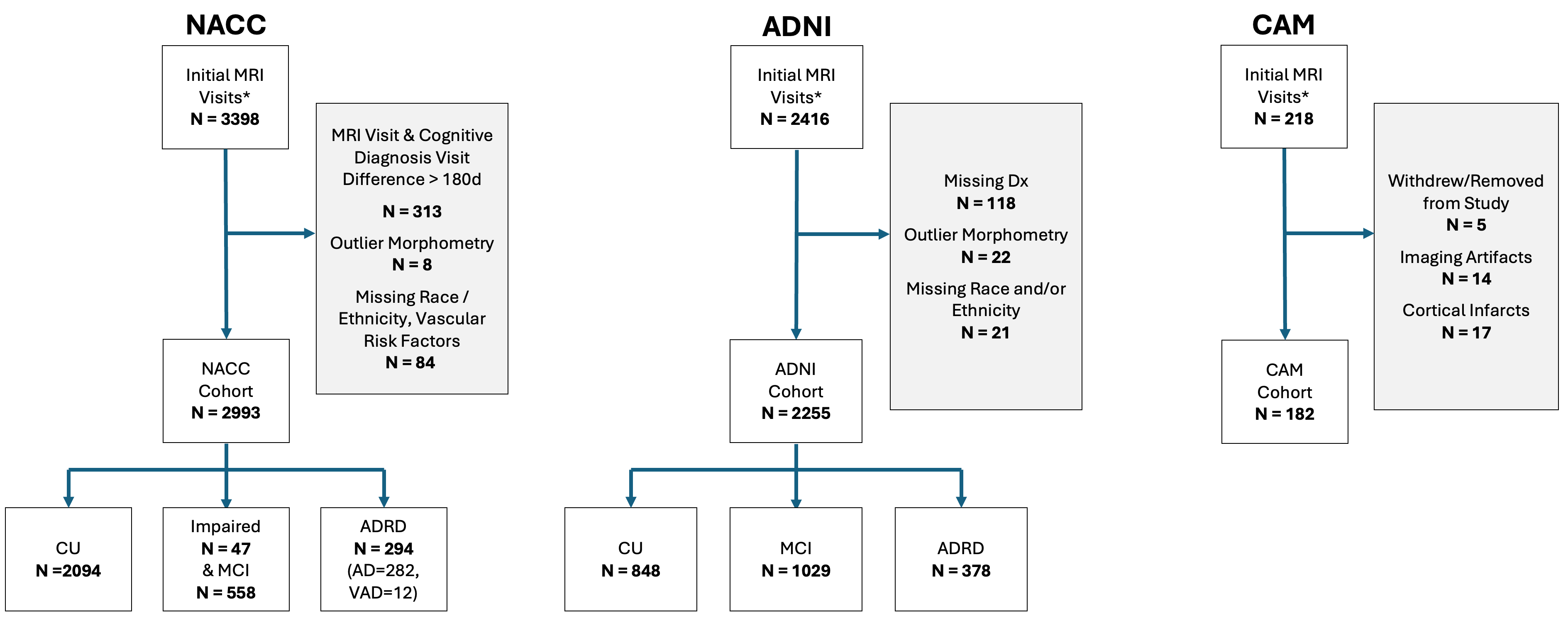

**Supplemental Figure 1. Baseline cross-sectional cohort selection flowcharts.** Flowcharts show participant selection for the baseline cross-sectional analyses within the National Alzheimer’s Coordinating Center (NACC), Alzheimer’s Disease Neuroimaging Initiative (ADNI), and Carotids and Minds (CAM) cohorts. Initial samples included participants with available baseline MRI. Participants were excluded for missing or discordant diagnosis data, outlier morphometry, demographic info, imaging artifacts, or cortical infarcts as applicable by cohort. Final analytic samples included 2,993 NACC participants, 2,255 ADNI participants, and 182 CAM participants. Diagnostic group distributions are shown for cognitively unimpaired (CU), mild cognitive impairment (MCI) / Impaired-not-MCI, and Alzheimer’s disease and related dementias (ADRD) (including Alzheimer’s disease [AD] and vascular dementia [VAD]). Cognitive status is not defined within the CAM cohort.

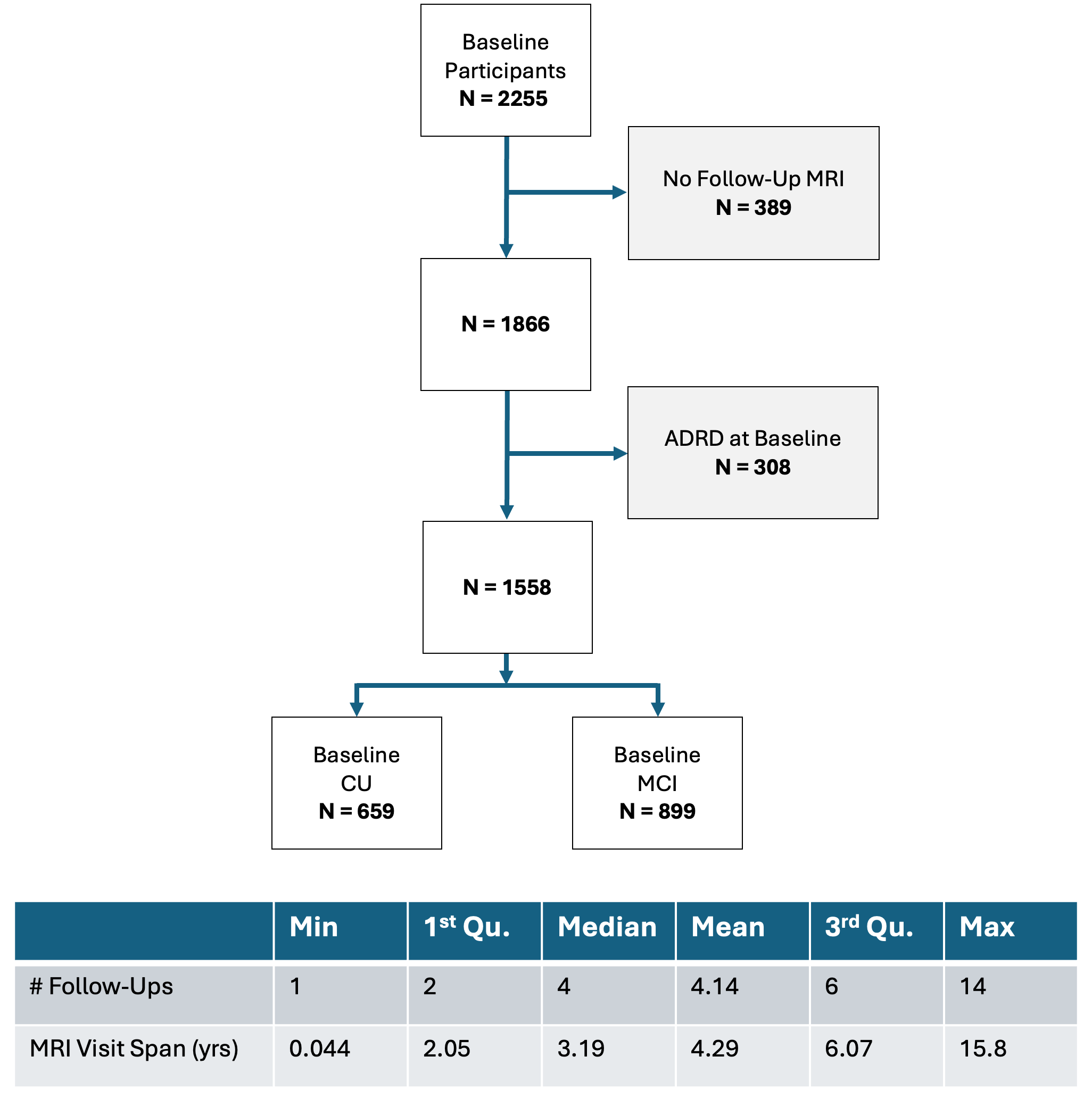

**Supplemental Figure 2. Longitudinal cohort selection flowchart.** Flowchart shows selection of ADNI participants included in longitudinal analyses of progression to ADRD. Of 2,255 baseline participants, 389 were excluded because they did not have follow-up MRI data. 308 participants with ADRD at baseline were also excluded, resulting in a final sample of 1,558 participants who were cognitively unimpaired (CU) or had mild cognitive impairment (MCI) at baseline. The table summarizes longitudinal follow-up characteristics, including number of follow-up visits and MRI visit span in years. Abbreviations: ADRD, Alzheimer’s disease and related dementias; CU, cognitively unimpaired; MCI, mild cognitive impairment.

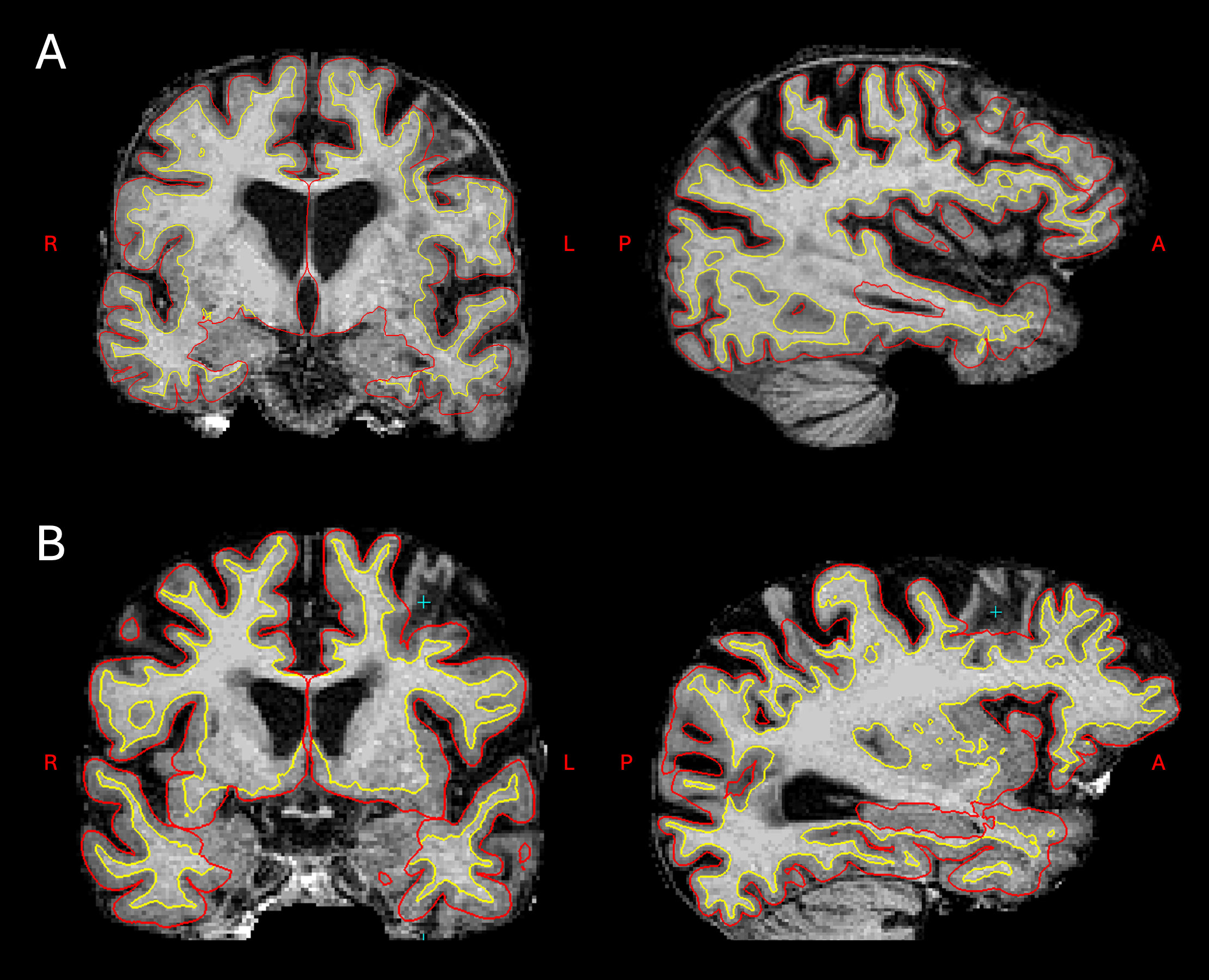

**Supplemental Figure 3. Segmentation errors and sample exclusions in Carotids and Minds cohort.** Examples of *FreeSurfer* segmentation errors due to cortical infarcts warranting exclusion from study analysis shown for two representative subjects in two orthogonal views (left: coronal, right: sagittal). (A) Segmentation error in remote infarcts of left frontal and occipital lobes. Failure to fully capture the left temporal lobe morphology can also be seen. (B) Segmentation error due to remote cortical infarct of left frontal lobe.

| **Baseline Characteristic** | **NACC**  **N = 2,993** | **ADNI**  **N = 2,255** | **CAM**  **N = 182** |
| --- | --- | --- | --- |
| **Age** | 71 (8) | 73 (7) | 72 (7) |
| **Sex** |  |  |  |
| Female | 1,897 (63%) | 1,063 (47%) | 85 (47%) |
| Male | 1,096 (37%) | 1,192 (53%) | 97 (53%) |
| **Education ^a^** | 16 (4) | 16 (3) | 15 (3) |
| **Race** |  |  |  |
| White | 2,231 (75%) | 2,013 (89%) | 176 (97%) |
| Black or African American | 601 (20%) | 158 (7.0%) | 1 (0.6%) |
| Asian | 71 (2.4%) | 55 (2.4%) | 4 (2.3%) |
| American Indian or Alaska Native | 1 (<0.1%) | 3 (0.1%) | 0 (0%) |
| Native Hawaiian / Pacific Islander | 42 (1.4%) | 1 (<0.1%) | 0 (0%) |
| More than one | NA | 25 (1.1%) | 0 (0%) |
| Other | 47 (1.6%) | 0 (0%) | 1 (0.6%) |
| **Ethnicity** |  |  |  |
| Hispanic/Latino | 268 (8.9%) | 91 (4.0%) | 25 (14%) |
| Not Hispanic/Latino | 2725 (91%) | 2,164 (96%) | 157 (86%) |
| **APOE ε4 ^b^** | 862 (38%) | 988 (46%) | 34 (27%) |
| **Hypertension** | 1,373 (46%) | 1,067 (47%) | 132 (73%) |
| **Diabetes** | 415 (14%) | 209 (9.3%) | 51 (28%) |
| **Hyperlipidemia** | 1,663 (56%) | 1,100 (49%) | 141 (77%) |
| **Smoking** | 495 (17%) | 521 (23%) | 96 (53%) |
| **Cognitive Status ^c^** |  |  |  |
| CU | 2,094 (70%) | 848 (38%) | NA |
| MCI | 605(20%) | 1,029 (46%) | NA |
| ADRD | 294 (9.8%) | 378 (17%) | NA |
| **Magnet Strength** |  |  |  |
| 3T | 2993 (100%) | 1,508 (67%) | 182 (100%) |
| 1.5T | 0 (0%) | 747 (33%) | 0 (%) |
| **Original/Processed Image** |  |  |  |
| Original | 2993 (100%) | 627 (28%) | 182 (100%) |
| Processed | 0 (0%) | 1,628 (72%) | 0 (0%) |

**Supplemental Table 1. Baseline characteristics of the cross-sectional cohorts.** Baseline demographic, clinical, diagnostic, and MRI acquisition characteristics are summarized for participants included in the cross-sectional analyses from NACC, ADNI, and CAM. Continuous variables are presented as mean (SD) and categorical variables as n(%). ^a^Education available for N=145 in CAM. ^b^*APOE* ε4 available for N=2276 in NACC, N=2171 in ADNI, N=125 in CAM. ^c^MCI category includes N=47 impaired-not-MCI participants, and ADRD category includes 12 non-AD vascular dementia participants in NACC. Abbreviations: ADNI, Alzheimer’s Disease Neuroimaging Initiative; ADRD, Alzheimer’s disease and related dementias; APOE, apolipoprotein E; CU, cognitively unimpaired; NACC, National Alzheimer’s Coordinating Center; MCI, mild cognitive impairment.

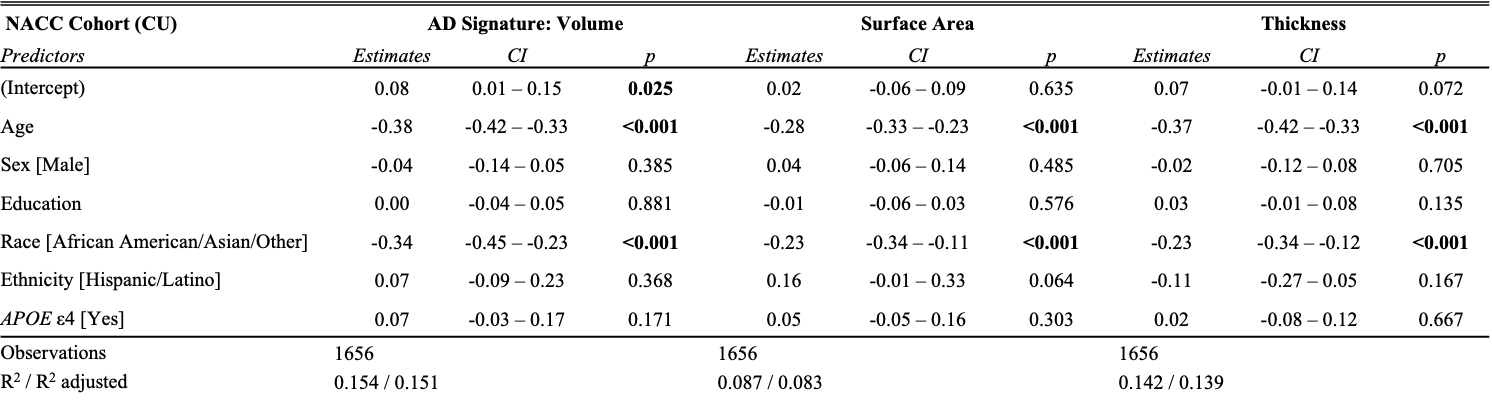

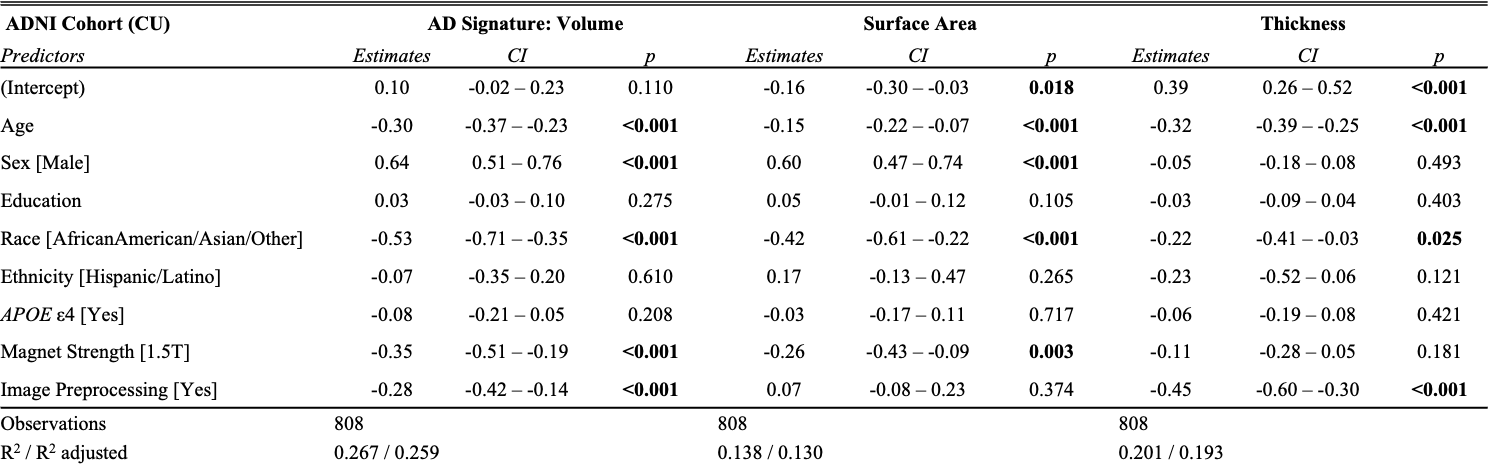
**Supplemental Figure 4. Association of AD-Signature morphometrics with demographic, genetic, and imaging characteristics in cognitively unimpaired participants.** Linear regression models show associations between demographic, genetic, and MRI acquisition covariates and AD-Signature cortical volume, surface area, and thickness among cognitively unimpaired participants in NACC (top row) and ADNI (bottom row). Beta estimates, 95% confidence intervals, and p-values are shown for each predictor. Models included age, sex, education years, race, ethnicity, and APOE e4 carrier status; ADNI models additionally included magnet strength and image processing status. Abbreviations: ADNI, Alzheimer’s Disease Neuroimaging Initiative; ADRD, Alzheimer’s disease and related dementias; APOE, apolipoprotein E; CI, confidence interval; CU, cognitively unimpaired; NACC, National Alzheimer’s Coordinating Center.

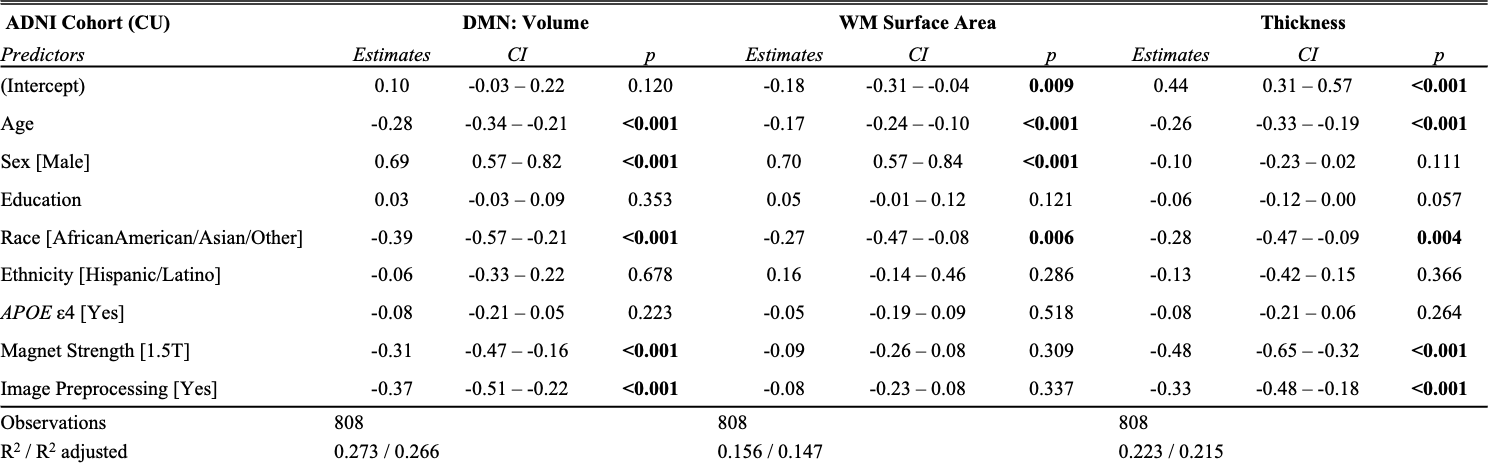

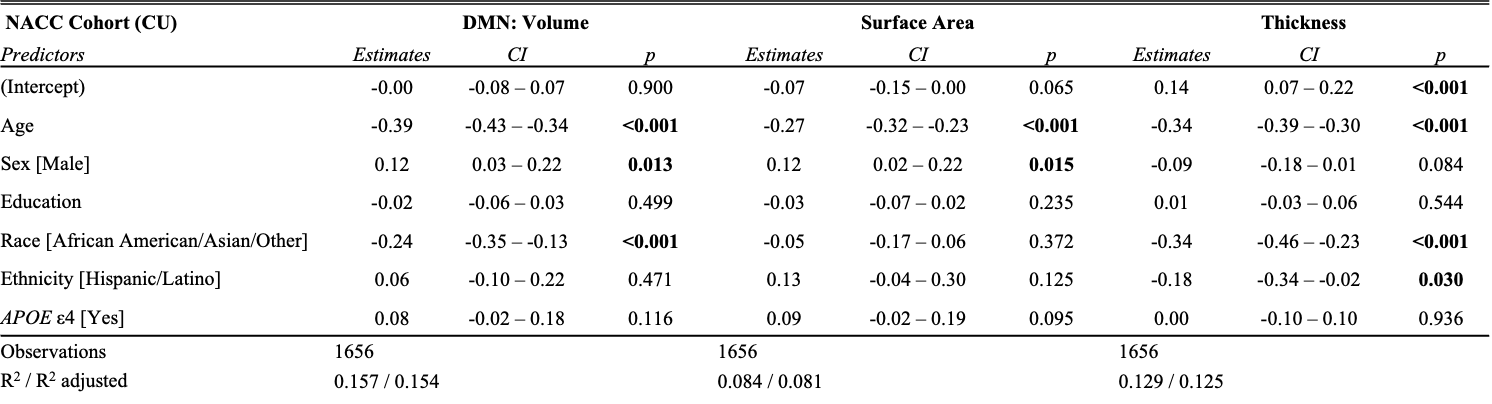

**Supplemental Figure 5. Association of DMN morphometrics with demographic, genetic, and imaging characteristics in cognitively unimpaired participants.** Linear regression models show associations between demographic, genetic, and MRI acquisition covariates and Default Mode Network (DMN) cortical volume, surface area, and thickness among CU participants in NACC (top row) and ADNI (bottom row). Beta estimates, 95% confidence intervals, and p-values are shown for each predictor. Models included age, sex, education years, race, ethnicity, and APOE e4 carrier status; ADNI models additionally included magnet strength and image processing status. Abbreviations: ADNI, Alzheimer’s Disease Neuroimaging Initiative; ADRD, Alzheimer’s disease and related dementias; APOE4, apolipoprotein E e4 allele; CI, confidence interval; CU, cognitively unimpaired; NACC, National Alzheimer’s Coordinating Center.

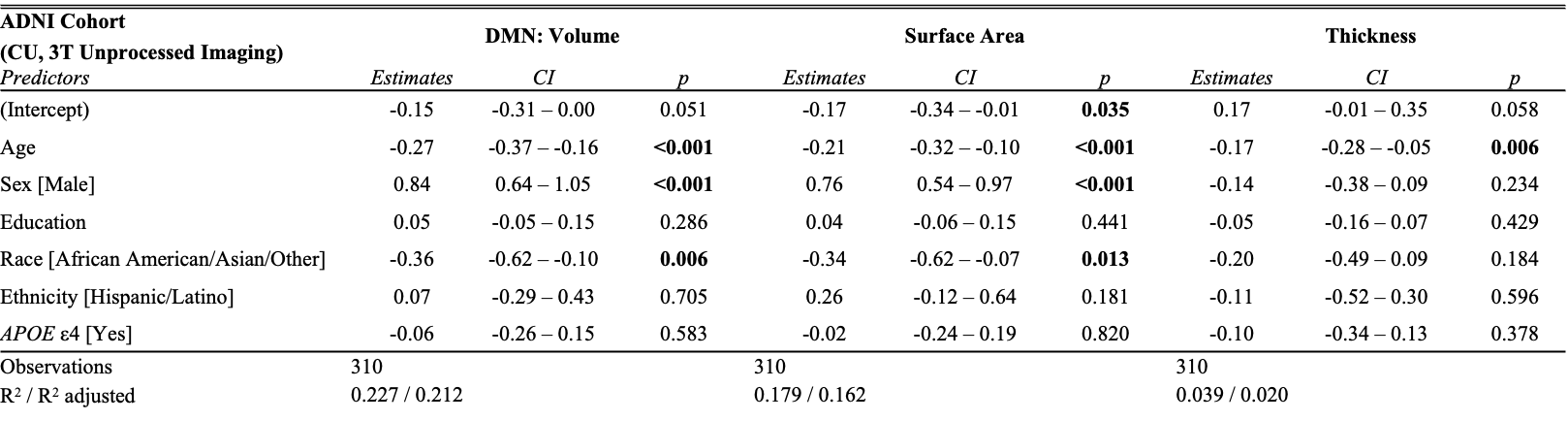

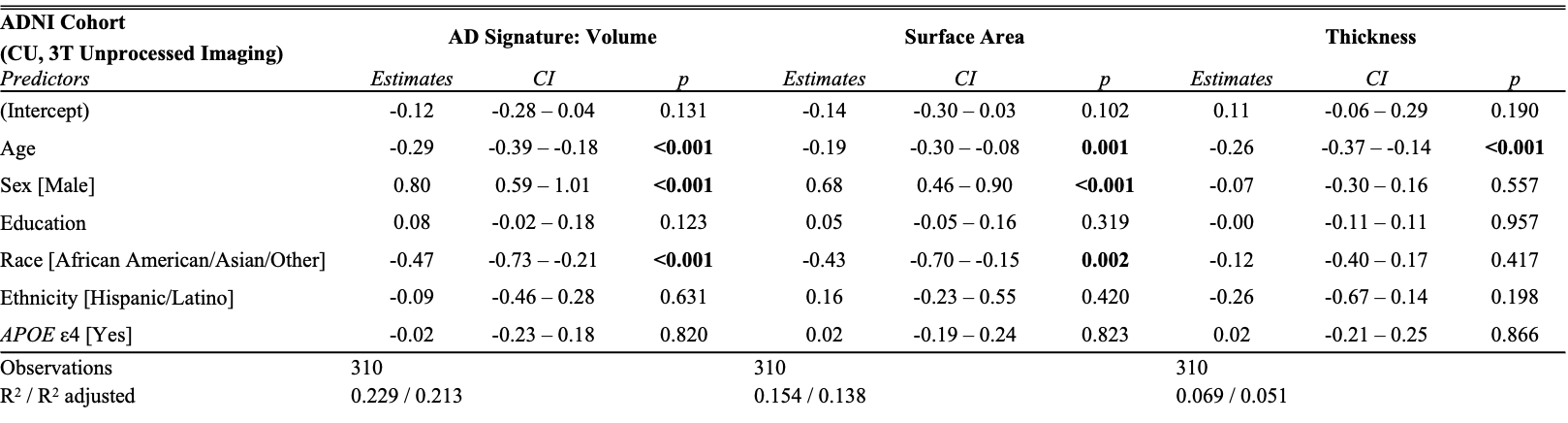

**Supplemental Figure 6. Sensitivity analysis of AD-Signature morphometrics among ADNI cognitively unimpaired participants with unprocessed 3T imaging.** Linear regression models show associations between demographic and genetic covariates and AD-Signature (top row) and DMN (bottom row) cortical volume, surface area, and thickness among CU participants after excluding those with pre-processed 1.5T imaging. Beta estimates, 95% confidence intervals, and p-values are shown for each predictor. Abbreviations: ADNI, Alzheimer’s Disease Neuroimaging Initiative; APOE, apolipoprotein E; CI, confidence interval; CU, cognitively unimpaired; DMN, Default Mode Network.

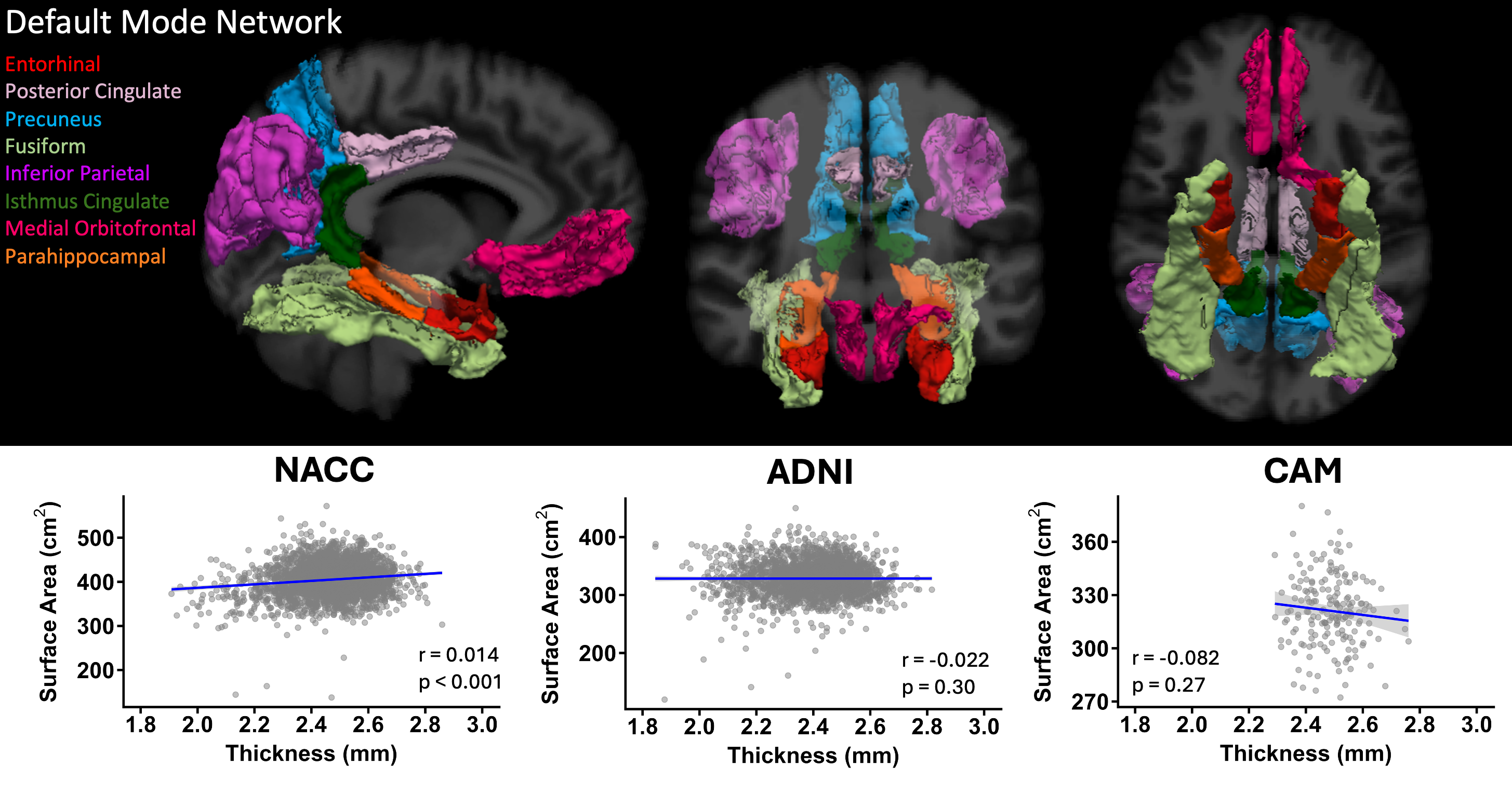

**Supplemental Figure 7. Default Mode Network composite regions and relationship between surface area and thickness across cohorts.** Top panels show the bilateral DMN-associated cortical regions included in the composite measure. Bottom panels show the relationship between DMN surface area and thickness in NACC, ADNI, and CAM cohorts. Shaded bands represent 95% confidence intervals around the fitted regression line. Spearman correlation coefficients and significance values are shown. Abbreviations: ADNI, Alzheimer’s Disease Neuroimaging Initiative; CAM, Carotids and Minds; DMN, Default Mode Network; NACC, National Alzheimer’s Coordinating Center.

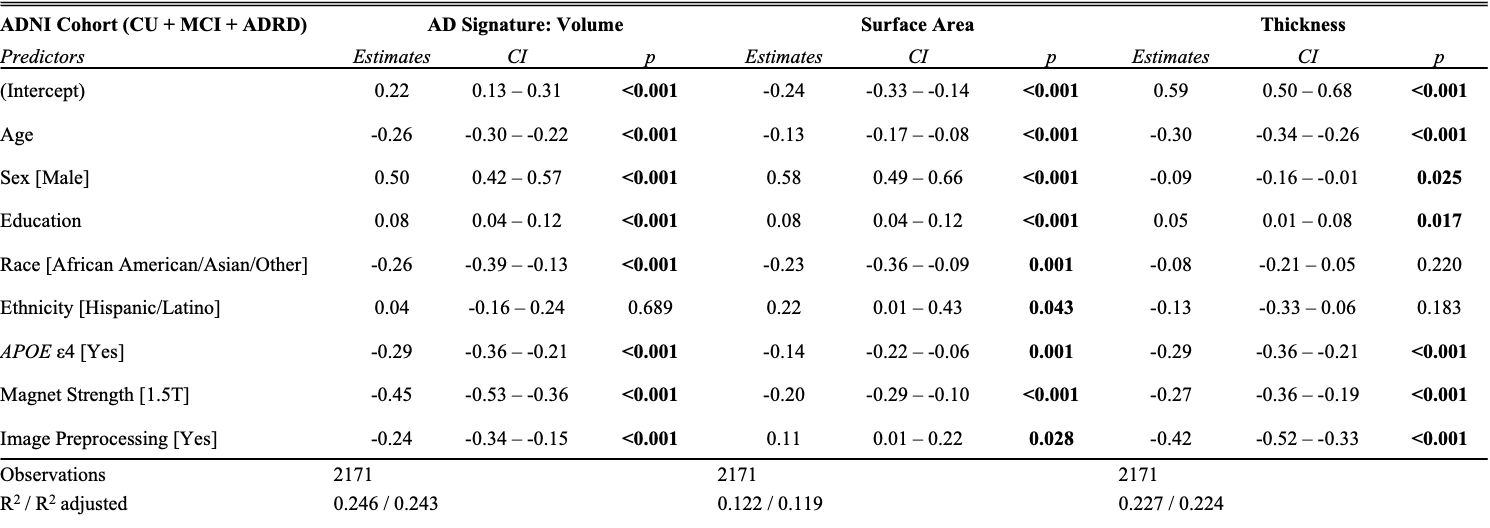

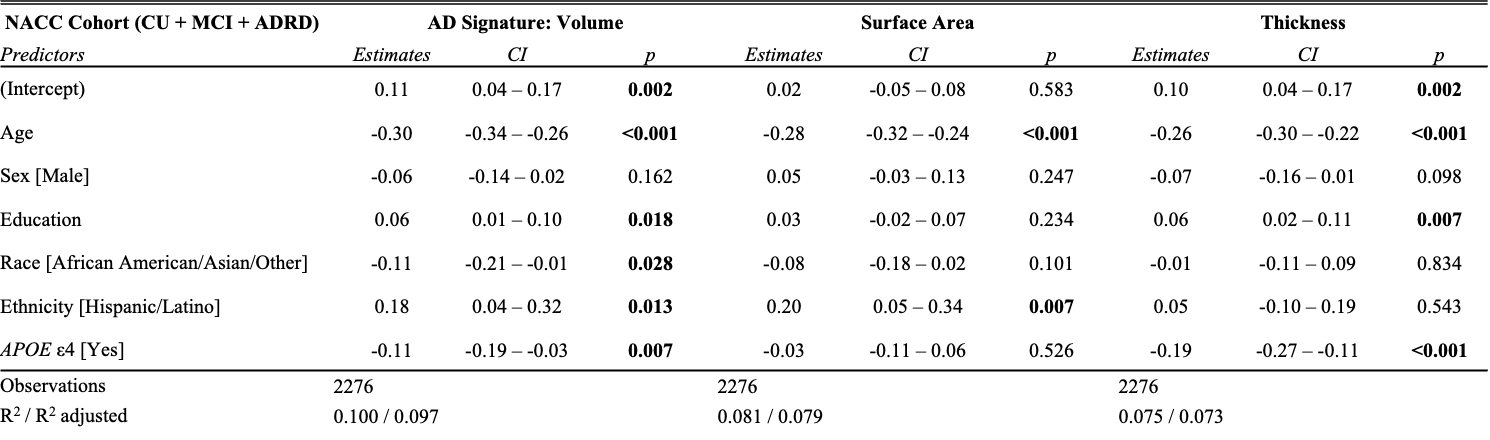
**Supplemental Figure 8. Full cohort NACC and ADNI AD-Signature morphometric associations with demographic, genetic, and imaging factors.** Linear regression models show associations between demographic, genetic, and MRI acquisition covariates and AD-Signature cortical volume, surface area, and thickness among CU, MCI, and ADRD participants in NACC (top) and ADNI (bottom). Beta estimates, 95% confidence intervals, and p-values are shown for each predictor. Models included age, sex, education years, race, ethnicity, and *APOE* e4 carrier status; ADNI models additionally included magnet strength and image processing status. Abbreviations: ADNI, Alzheimer’s Disease Neuroimaging Initiative; ADRD, Alzheimer’s disease and related dementias; APOE, apolipoprotein E; CI, confidence interval; CU, cognitively unimpaired; Dem., dementia; MCI, mild cognitive impairment; NACC, National Alzheimer’s Coordinating Center.

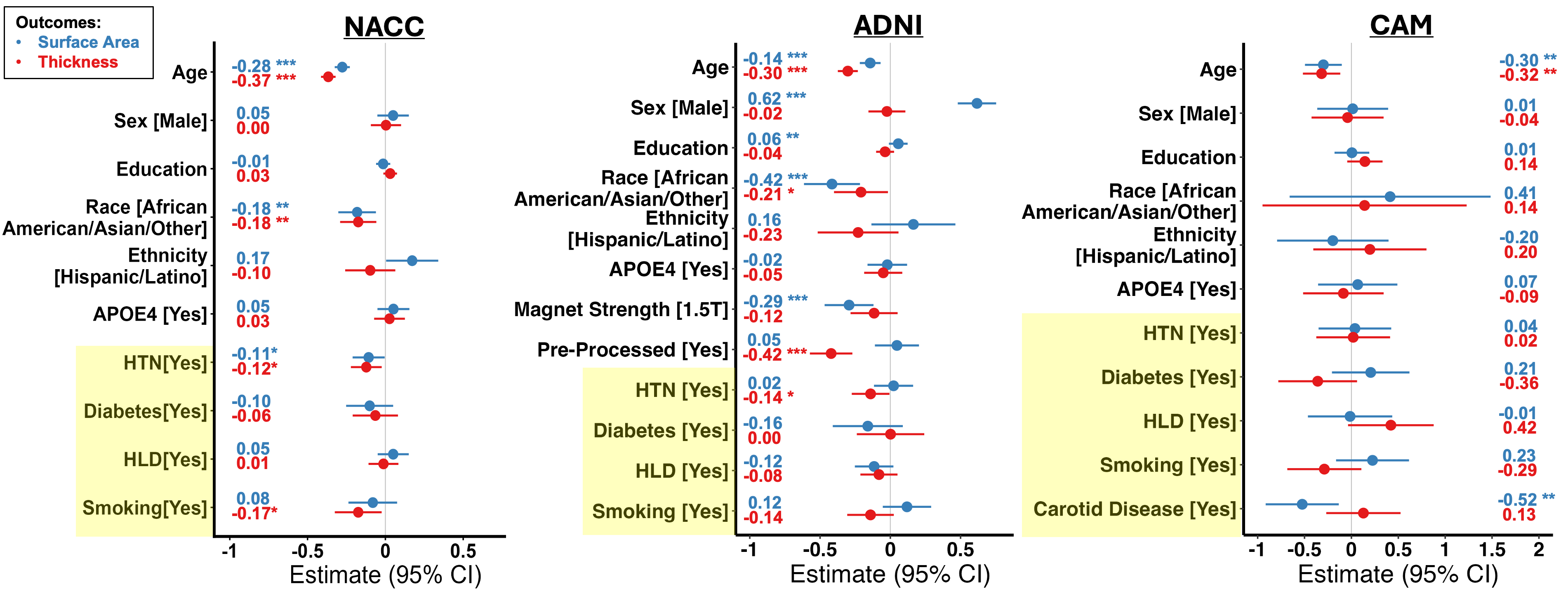

**Supplemental Figure 9. AD-Signature surface area and thickness associations with vascular risk factors and carotid disease in cognitively unimpaired participants.** Forest plots show adjusted associations of vascular risk factors with AD-Signature surface area (blue) and thickness (red) in CU participants. Models are shown for all three cohorts (from left to right): NACC, ADNI, and CAM. Beta estimates and 95% confidence intervals are shown for each predictor. Models included demographic and genetic covariates across cohorts, with imaging characteristics included in ADNI. Carotid disease was also assessed in CAM. Abbreviations: ADNI, Alzheimer’s Disease Neuroimaging Initiative; APOE4, apolipoprotein E e4 allele; CAM, Carotids and Minds; CU, cognitively unimpaired; HLD, hyperlipidemia; HTN, hypertension; NACC, National Alzheimer’s Coordinating Center.

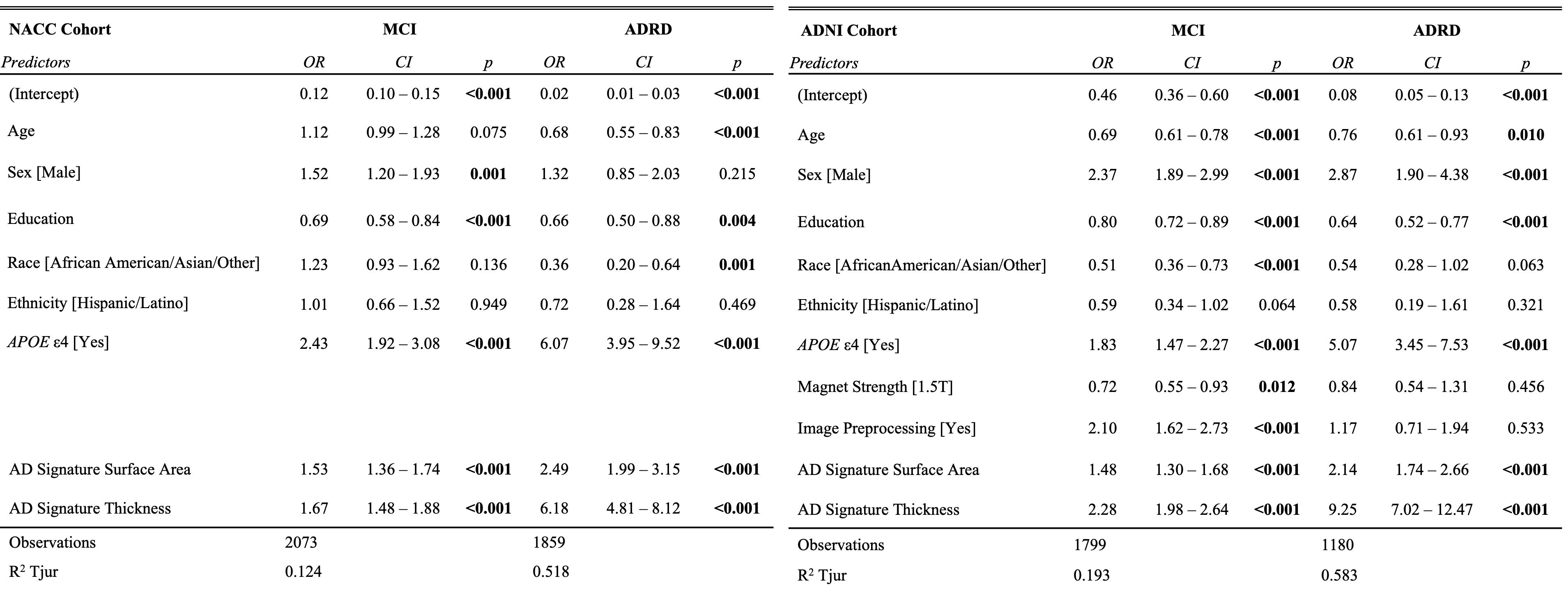

**Supplemental Figure 10. Independent associations of AD-Signature surface area and thickness with cognitive diagnosis in NACC and ADNI.** Multivariable logistic regression models show the independent associations of AD-Signature cortical surface area and cortical thickness with cognitive diagnosis in NACC (left) and ADNI (right) cohorts. CU participants served as the reference group. Odds ratios, 95% confidence intervals, and p-values are shown for MCI and ADRD in NACC and ADNI. Models included age, sex, education years, race, ethnicity, APOE e4 carrier status, AD-Signature cortical surface area and thickness; ADNI model additionally included magnet strength and image processing status. Abbreviations: ADNI, Alzheimer’s Disease Neuroimaging Initiative; ADRD, Alzheimer’s disease and related dementias; APOE, apolipoprotein E; CI, confidence interval; CU, cognitively unimpaired; NACC, National Alzheimer’s Coordinating Center; MCI, mild cognitive impairment; OR, odds ratio.

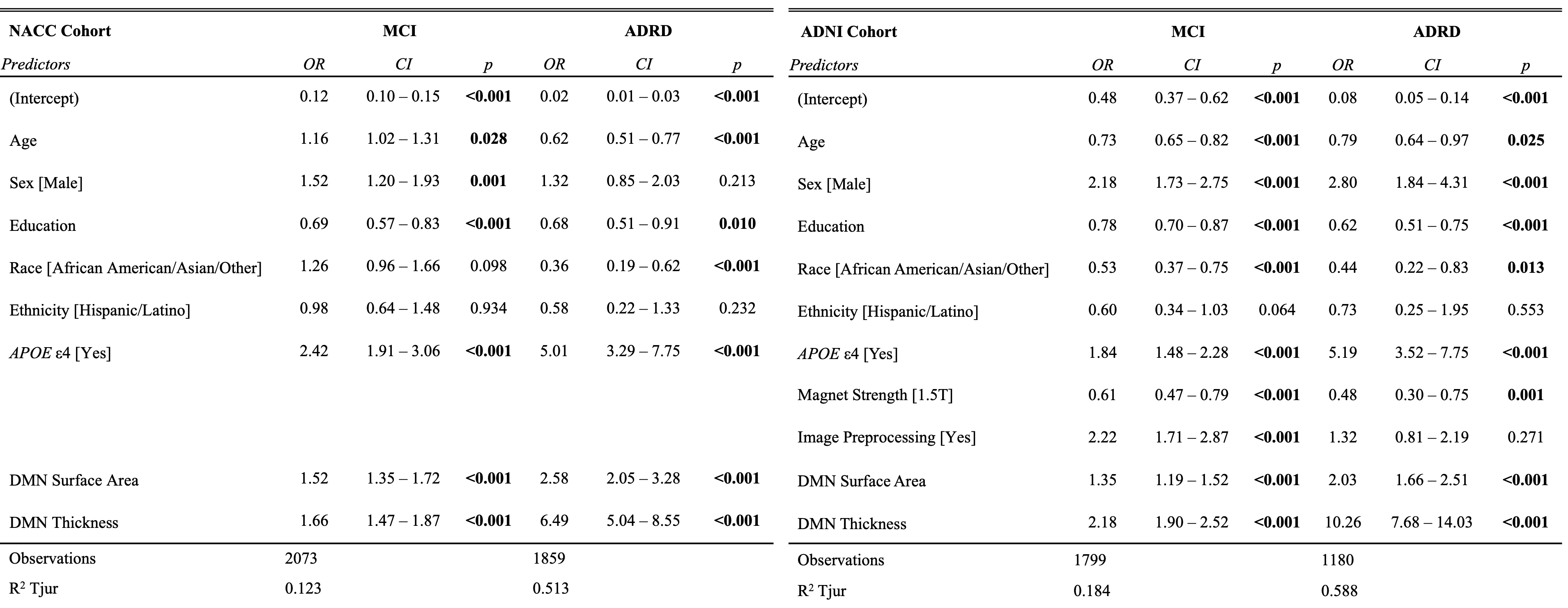

**Supplemental Figure 11. Independent associations of Default Mode Network surface area and thickness with cognitive diagnosis in NACC and ADNI.** Multivariable logistic regression models show the independent associations of DMN cortical surface area and cortical thickness with cognitive diagnosis in NACC (left) and ADNI (right) cohorts. CU participants served as the reference group. Odds ratios, 95% confidence intervals, and p-values are shown for MCI and ADRD in NACC and ADNI. Models included age, sex, education years, race, ethnicity, APOE e4 carrier status, AD-Signature cortical surface area and thickness; ADNI model additionally included magnet strength and image processing status. Abbreviations: ADNI, Alzheimer’s Disease Neuroimaging Initiative; ADRD, Alzheimer’s disease and related dementias; APOE, apolipoprotein E; CI, confidence interval; CU, cognitively unimpaired; DMN, Default Mode Network; NACC, National Alzheimer’s Coordinating Center; MCI, mild cognitive impairment; OR, odds ratio.

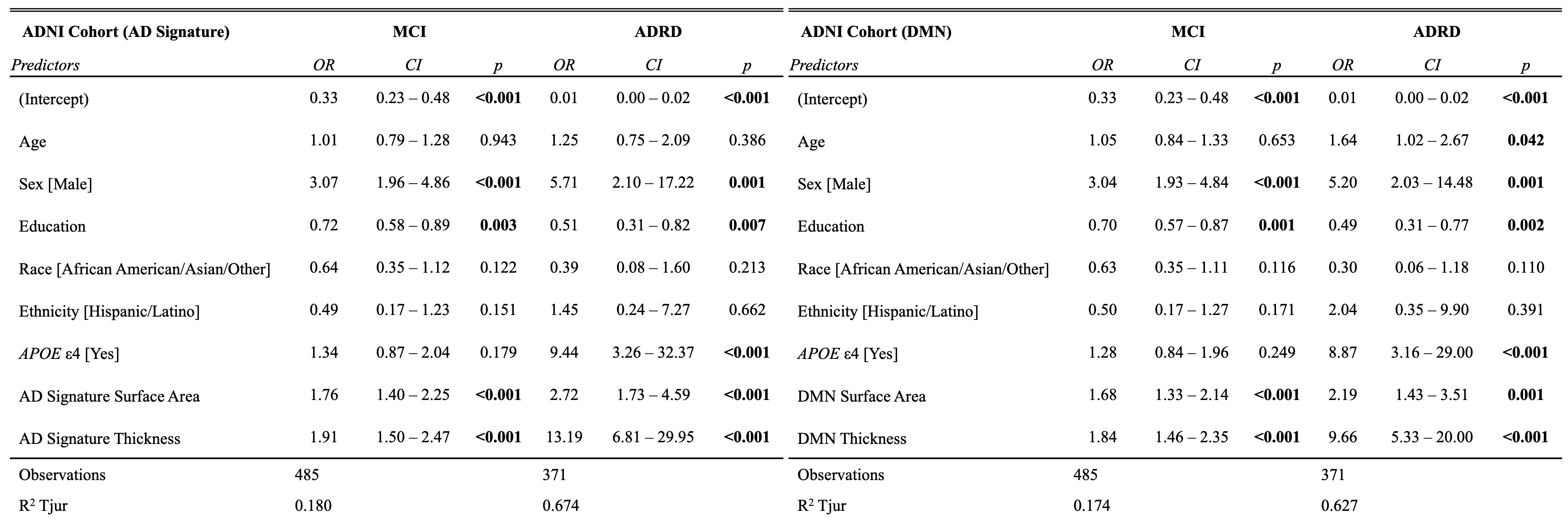

**Supplemental Figure 12. Sensitivity analysis of independent associations between cortical surface area, cortical thickness, and cognitive diagnosis in ADNI unprocessed 3T imaging.** Logistic regression models show the independent associations of AD-Signature (left) and DMN (right) cortical surface area and thickness with MCI and ADRD in a restricted ADNI sample excluding participants with preprocessed 1.5T imaging. CU participants served as the reference group. Odds ratios, 95% confidence intervals, and p-values are shown for each predictor. Abbreviations: ADNI, Alzheimer’s Disease Neuroimaging Initiative; ADRD, Alzheimer’s disease and related dementias; APOE, apolipoprotein E; CI, confidence interval; CU, cognitively unimpaired; DMN, Default Mode Network; MCI, mild cognitive impairment; OR, odds ratio.

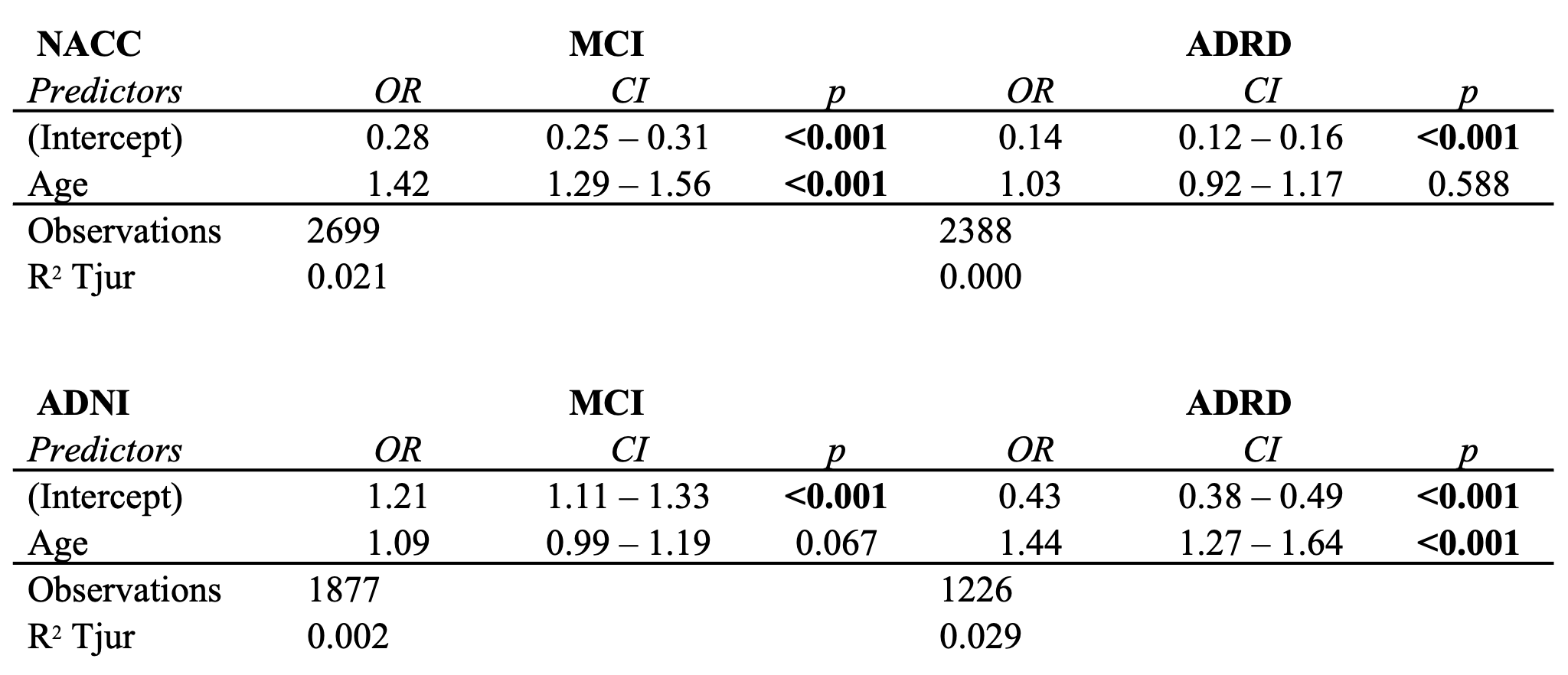

**Supplemental Figure 13. Age-only associations with MCI and AD/dementia in NACC and ADNI.** Unadjusted logistic regression models show the association between age and cognitive diagnosis in NACC (top) and ADNI (bottom) without additional demographics, genetic, MRI acquisition, or morphometric covariates. CU participants served as the reference group. Odds ratios, 95% confidence intervals, and p-values are shown. In age-only models, older age was associated with greater odds of MCI and ADRD, though not statistically significant for ADRD in NACC or MCI in ADNI. These analyses were performed to evaluate whether inverse age associations observed in fully adjusted diagnostic models (Supplemental Figures 11-12) reflected covariate adjustment rather than the association between age and diagnosis. Abbreviations: ADNI, Alzheimer’s Disease Neuroimaging Initiative; ADRD, Alzheimer’s disease and related dementias; CI, confidence interval; CU, cognitively unimpaired; MCI, mild cognitive impairment; NACC, National Alzheimer’s Coordinating Center; OR, odds ratio.

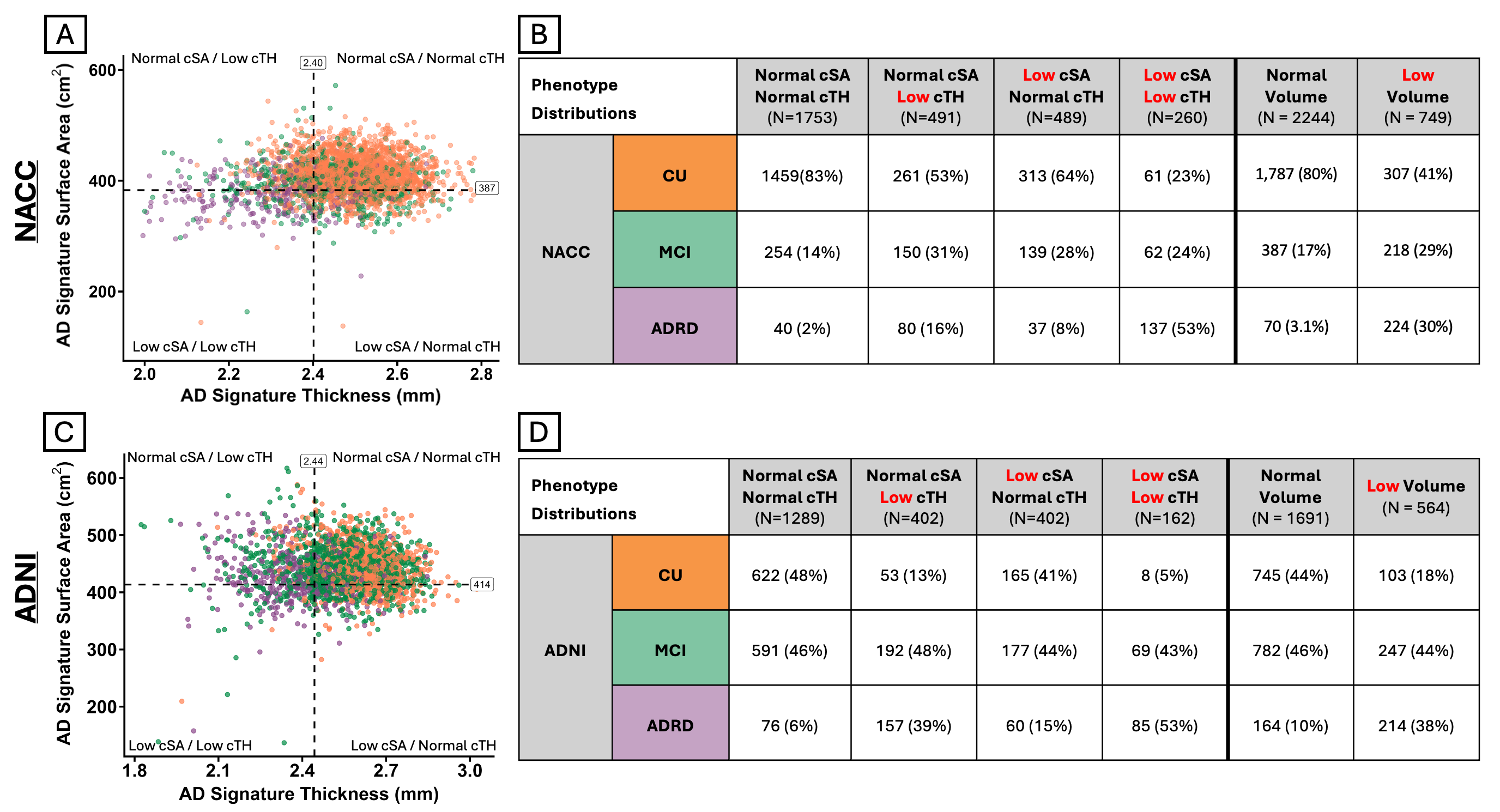

**Supplemental Figure 14. AD-Signature surface area/thickness phenotypes and diagnostic distributions in NACC and ADNI.** Scatterplots show the distribution of AD-Signature cortical surface area (cSA) and thickness (cTH) in the NACC (A) and ADNI (C) cohorts. Dashed lines indicate cohort-specific lower-quartile cut points used to define high versus low cSA and cTH**,** generating four morphometric phenotypes: high cSA/high cTH, high cSA/low cTH, low cSA/high cTH, low cSA/low cTH. Corresponding tables show the diagnostic composition of each cSA/cTH and volume phenotype in NACC (B) and ADNI (D). Across both cohorts, the low cSA/low cTH phenotype had the highest proportion of ADRD participants, whereas the high cSA/high cTH phenotype had the largest proportion of CU participants. Volume-based high/low groups (defined by lower quartile cut point) are shown for comparison. Abbreviations: ADNI, Alzheimer’s Disease Neuroimaging Initiative; ADRD, Alzheimer’s disease and related dementias; cSA, cortical surface area; cTH, cortical thickness; CU, cognitively unimpaired; MCI, mild cognitive impairment; NACC, National Alzheimer’s Coordinating Center.

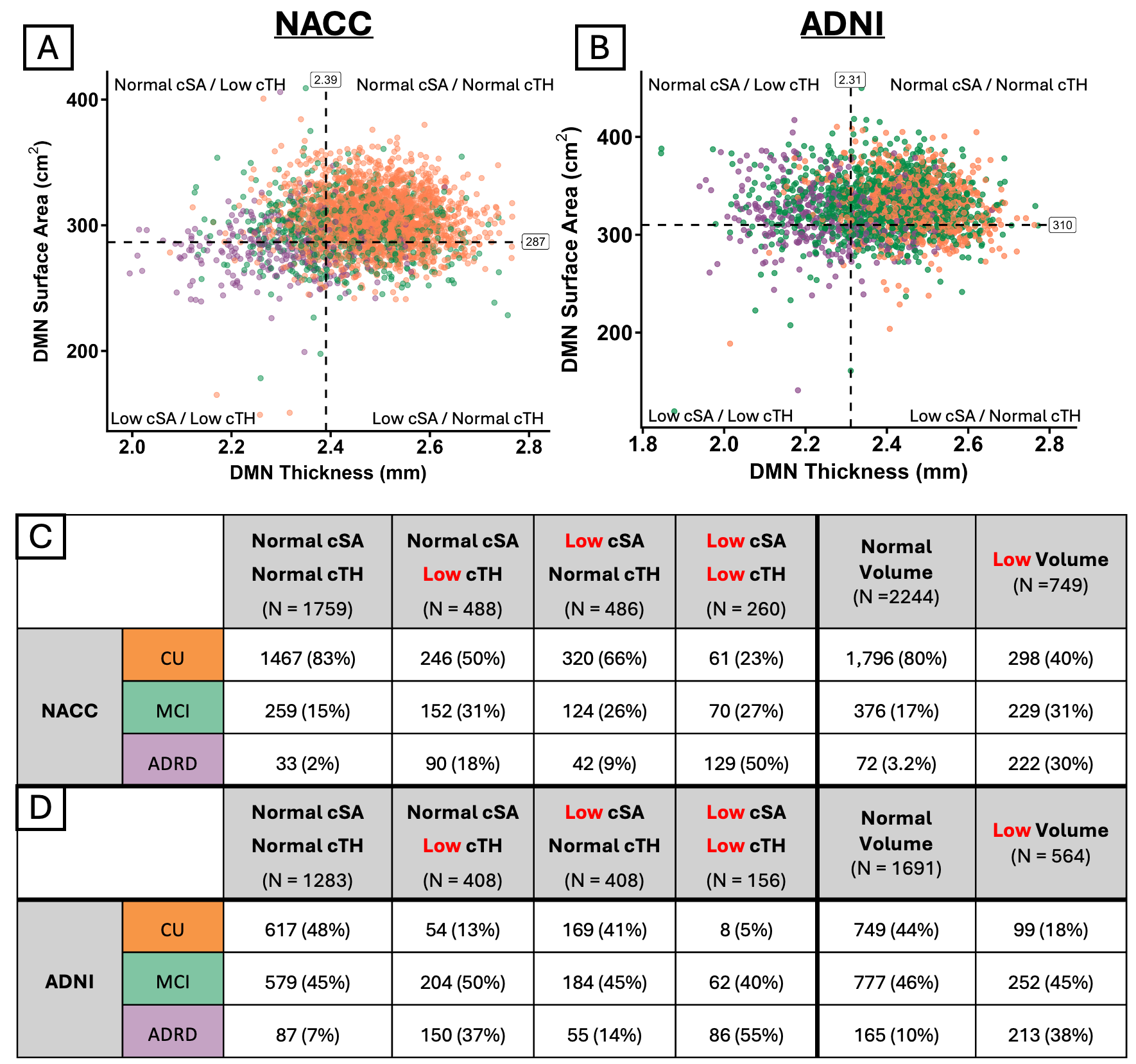

**Supplemental Figure 15. DMN surface area/thickness phenotypes and diagnostic distributions in NACC and ADNI.** Scatterplots show the distribution of DMN cortical surface area (cSA) and thickness (cTH) in the NACC (A) and ADNI (B) cohorts. Dashed lines indicate cohort-specific lower-quartile cut points used to define high versus low cSA and cTH**,** generating four morphometric phenotypes: high cSA/high cTH, high cSA/low cTH, low cSA/high cTH, low cSA/low cTH. Corresponding tables show the diagnostic composition of each cSA/cTH and volume phenotype in NACC (C) and ADNI (D). Across both cohorts, the low cSA/low cTH phenotype had the highest proportion of ADRD participants, whereas the high cSA/high cTH phenotype had the largest proportion of CU participants. Volume-based high/low groups (defined by lower quartile cut point) are shown for comparison. Abbreviations: ADNI, Alzheimer’s Disease Neuroimaging Initiative; ADRD, Alzheimer’s disease and related dementias; cSA, cortical surface area; cTH, cortical thickness; CU, cognitively unimpaired; DMN, Default Mode Network; MCI, mild cognitive impairment; NACC, National Alzheimer’s Coordinating Center.

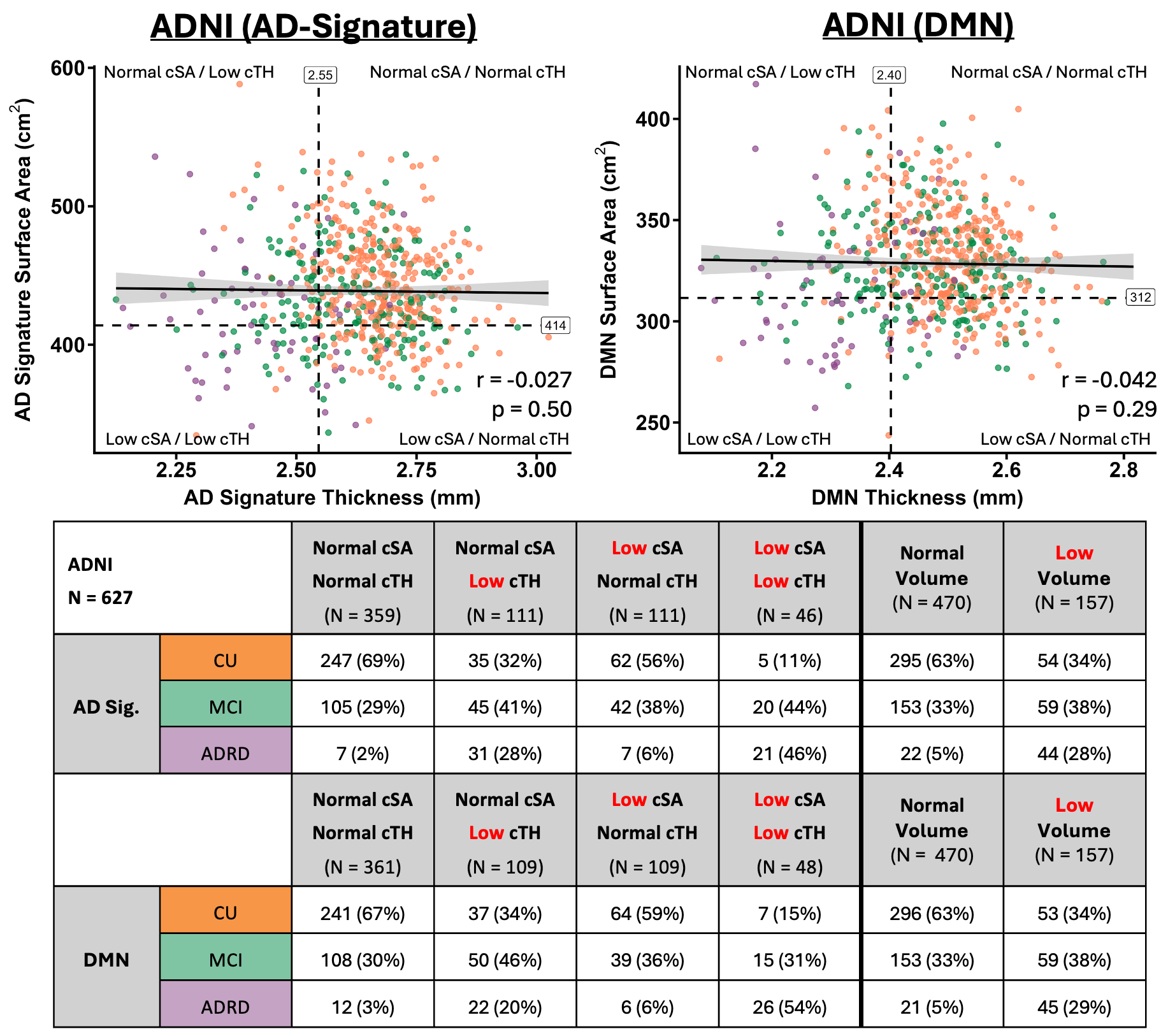

**Supplemental Figure 16. Sensitivity analysis of ADNI surface area/thickness phenotypes in participants with unprocessed 3T imaging.** Scatterplots show the distribution of cortical surface area (cSA) and thickness (cTH) in ADNI participants having unprocessed 3T imaging for AD-Signature (A) and DMN (B). Dashed lines indicate cohort-specific lower-quartile cut points used to define high versus low cSA and cTH**,** generating four morphometric phenotypes: high cSA/high cTH, high cSA/low cTH, low cSA/high cTH, low cSA/low cTH. Spearman correlation coefficients and p-values are shown. Corresponding tables show the diagnostic composition of each cSA/cTH and volume phenotype for AD-Signature (C) and DMN (D). Across both cohorts, the low cSA/low cTH phenotype had the highest proportion of ADRD participants, whereas the high cSA/high cTH phenotype had the largest proportion of CU participants. Volume-based high/low groups (defined by lower quartile cut point) are shown for comparison. Abbreviations: ADNI, Alzheimer’s Disease Neuroimaging Initiative; ADRD, Alzheimer’s disease and related dementias; cSA, cortical surface area; cTH, cortical thickness; CU, cognitively unimpaired; DMN, Default Mode Network; MCI, mild cognitive impairment.

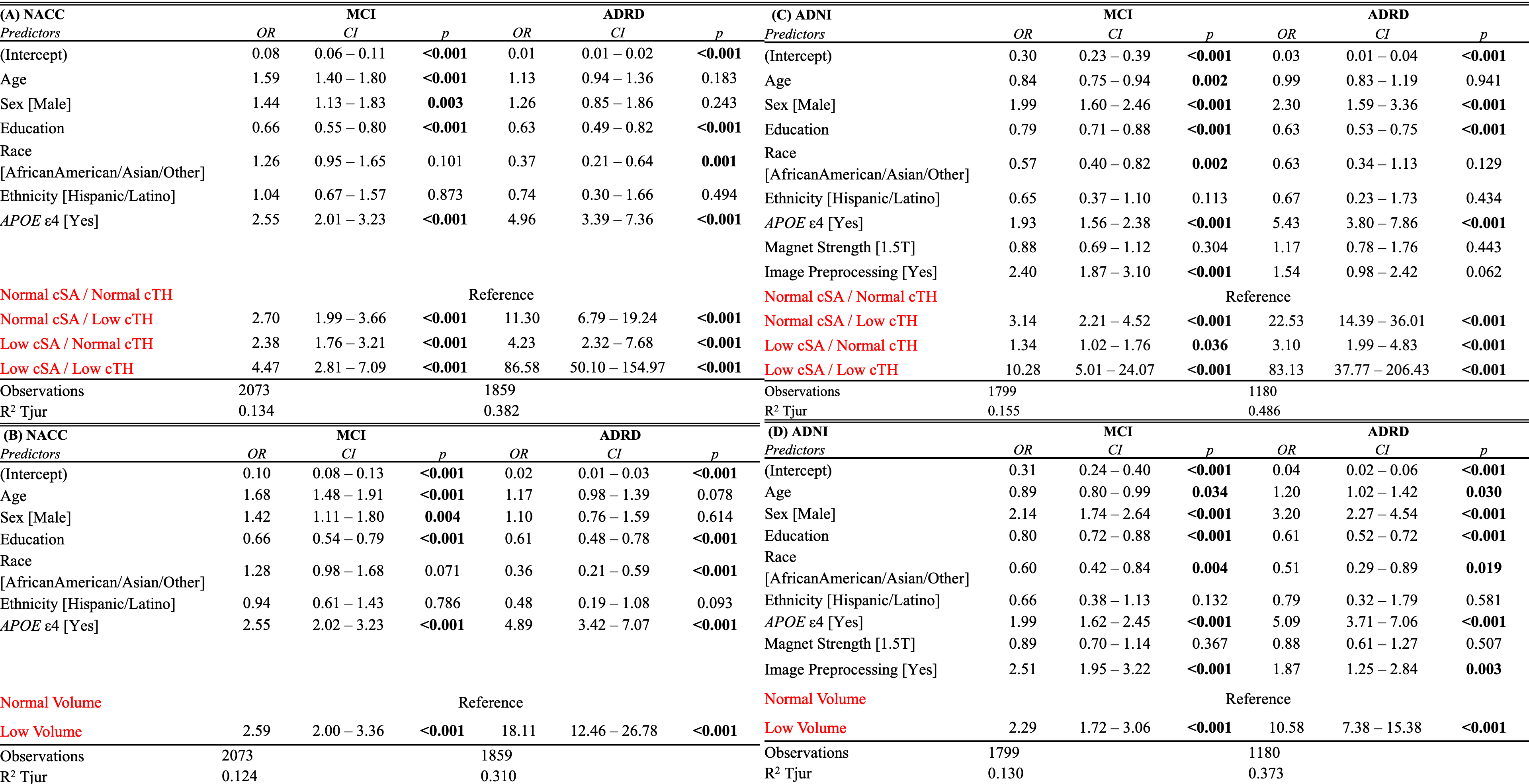

**Supplemental Figure 17. Full models for AD-Signature surface area/thickness phenotypes and volume-defined groups in NACC and ADNI.** Logistic regression models show associations of AD-Signature cSA/cTH phenotypes and AD-Signature volume-defined groups with cognitive diagnosis in NACC (A-B) and ADNI (C-D). Models were fit separately for cSA/cTH phenotypes and volume-defined groups. CU participants served as the reference group. Odds ratios, 95% confidence intervals, and p-values are shown for MCI and ADRD in NACC and ADNI. Models included age, sex, education years, race, ethnicity, APOE e4 carrier status, and the morphometric phenotype or volume group; ADNI models additionally included magnet strength and image processing status. Across both cohorts, low cSA/low cTH was associated with the greatest odds of MCI and ADRD. Low volume was also associated with greater odds of MCI and ADRD. Abbreviations: ADNI, Alzheimer’s Disease Neuroimaging Initiative; ADRD, Alzheimer’s disease and related dementias; APOE, apolipoprotein E; CI, confidence interval; cSA, cortical surface area; cTH, cortical thickness; CU, cognitively unimpaired; MCI, mild cognitive impairment; NACC, National Alzheimer’s Coordinating Center; OR, odds ratio.

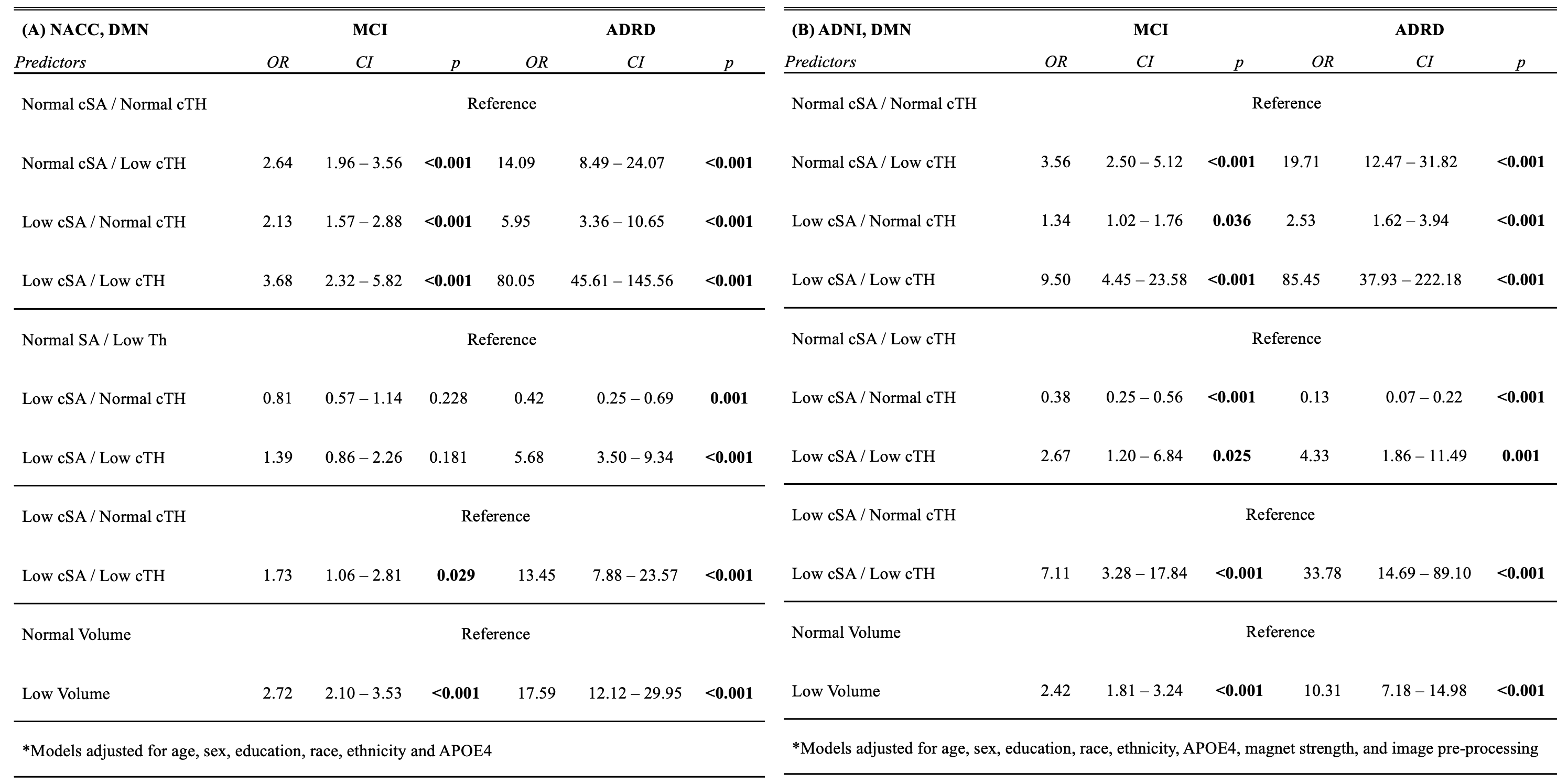

**Supplemental Figure 18. Diagnostic associations of DMN surface area/thickness phenotypes and volume-defined groups in NACC and ADNI.** Logistic regression models show associations of DMN cSA/cTH phenotypes and volume-defined groups with cognitive diagnosis in NACC and ADNI. CU participants served as the reference group. For cSA/cTH models, the high cSA/high cTH phenotype served as the primary reference group, with additional pairwise comparisons shown to evaluate whether the combined low cSA/low cTH phenotype was associated with greater odds than the intermediate phenotypes. Odds ratios, 95% confidence intervals, and p-values are shown for MCI and ADRD in NACC and ADNI. Models were adjusted for age, sex, education years, race, ethnicity, and APOE e4 carrier status; ADNI models additionally included magnet strength and image preprocessing status. Across both cohorts, low cSA/low cTH was associated with the greatest odds of ADRD, exceeding odds observed for either low cSA or low cTH alone. Abbreviations: ADNI, Alzheimer’s Disease Neuroimaging Initiative; ADRD, Alzheimer’s disease and related dementias; APOE, apolipoprotein E; CI, confidence interval; cSA, cortical surface area; cTH, cortical thickness; CU, cognitively unimpaired; DMN, Default Mode Network; MCI, mild cognitive impairment; NACC, National Alzheimer’s Coordinating Center; OR, odds ratio.

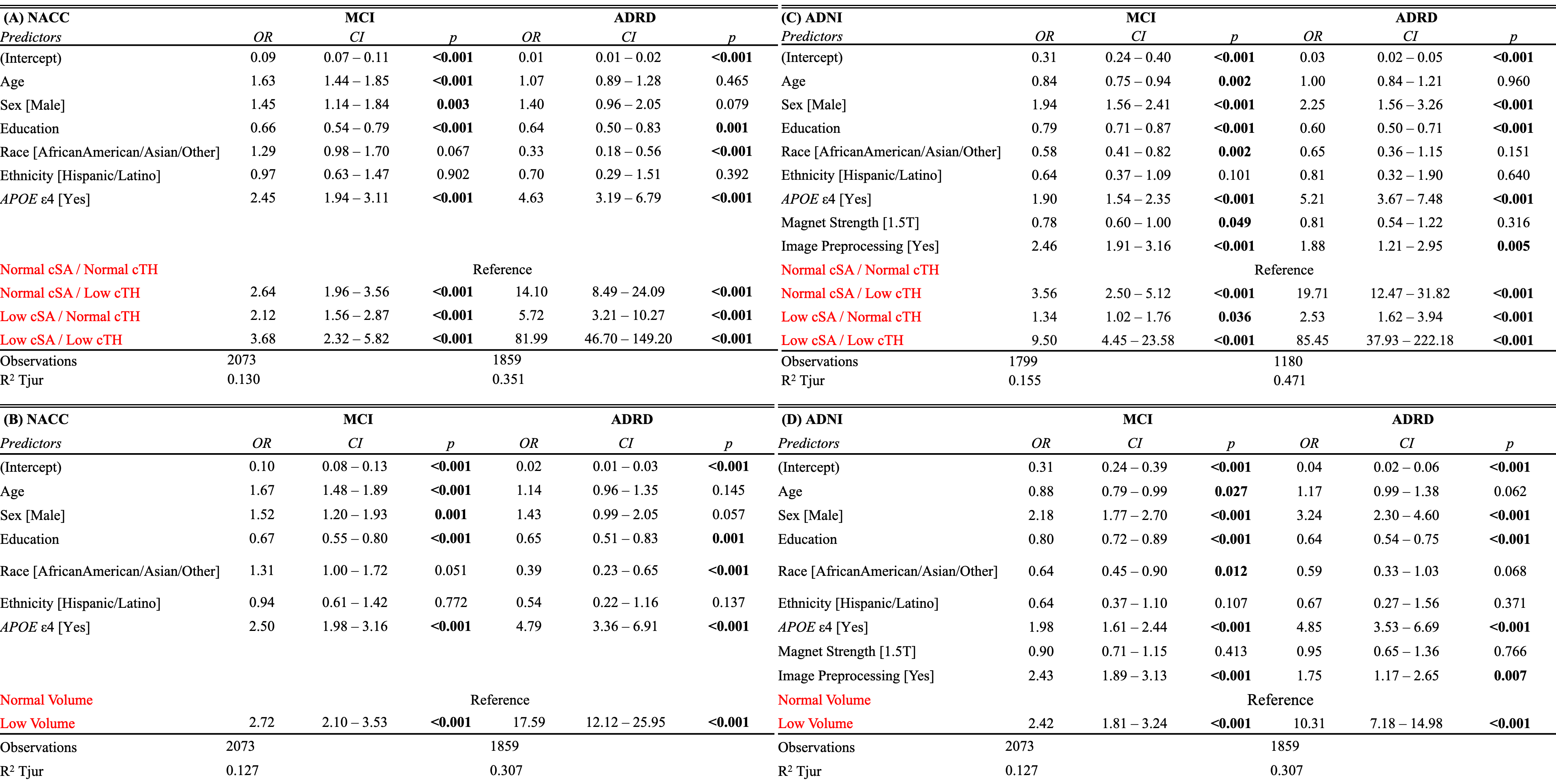

**Supplemental Figure 19. Full models for DMN surface area/thickness phenotypes and volume-defined groups in NACC and ADNI.** Logistic regression models show associations of DMN cSA/cTH phenotypes and AD-Signature volume-defined groups with cognitive diagnosis in NACC (A-B) and ADNI (C-D). Models were fit separately for cSA/cTH phenotypes and volume-defined groups. CU participants served as the reference group. Odds ratios, 95% confidence intervals, and p-values are shown for MCI and ADRD in NACC and ADNI. Models included age, sex, education years, race, ethnicity, APOE e4 carrier status, and the morphometric phenotype or volume group; ADNI models additionally included magnet strength and image processing status. Across both cohorts, low cSA/low cTH was associated with the greatest odds of MCI and ADRD. Low volume was also associated with greater odds of MCI and ADRD. Abbreviations: ADNI, Alzheimer’s Disease Neuroimaging Initiative; ADRD, Alzheimer’s disease and related dementias; APOE, apolipoprotein E; CI, confidence interval; cSA, cortical surface area; cTH, cortical thickness; CU, cognitively unimpaired; DMN, Default Mode Network; MCI, mild cognitive impairment; NACC, National Alzheimer’s Coordinating Center; OR, odds ratio.

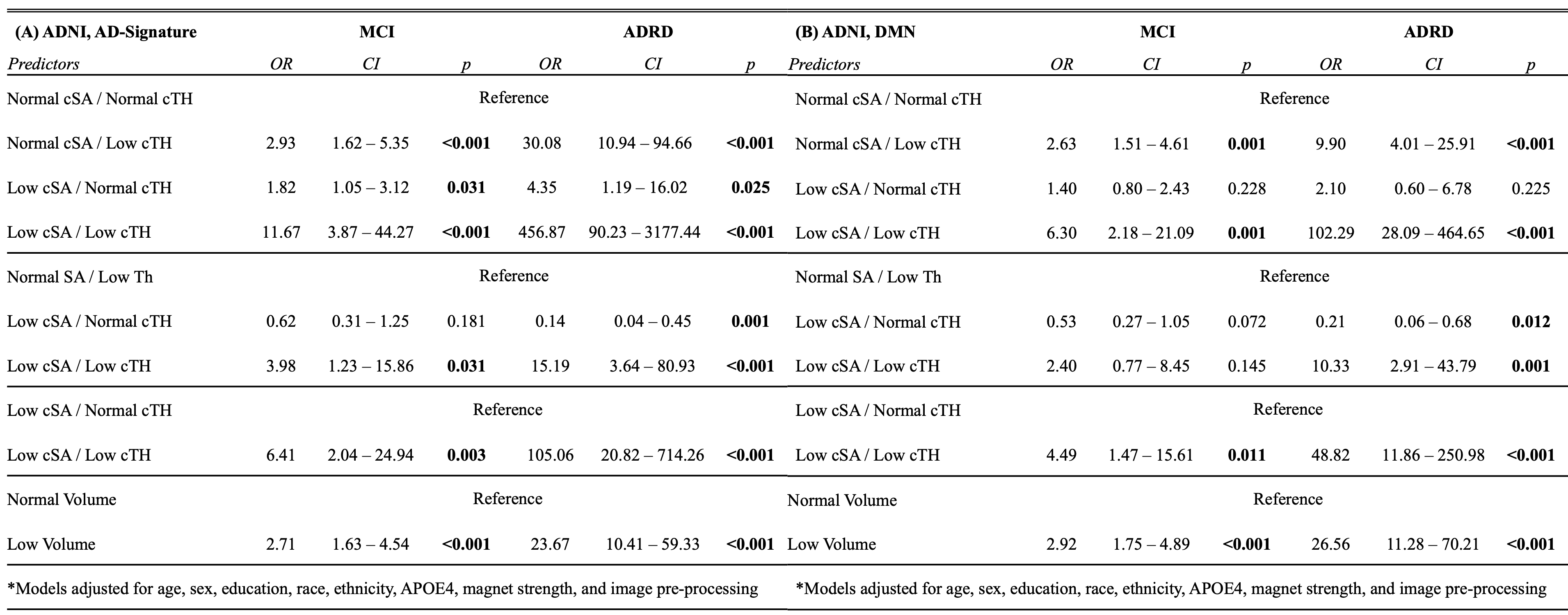

**Supplemental Figure 20. Sensitivity analysis of diagnostic associations with AD-Signature and DMN surface area/thickness phenotypes and volume-defined groups in participants with 3T unprocessed imaging.** Logistic regression models show associations of AD-Signature (A) and DMN (B) cSA/cTH phenotypes and volume-defined groups with cognitive diagnosis in ADNI, excluding participants with 1.5T preprocessed imaging. CU participants served as the reference group. For cSA/cTH models, the high cSA/high cTH phenotype served as the primary reference group, with additional pairwise comparisons shown to evaluate whether the combined low cSA/low cTH phenotype was associated with greater odds than the intermediate phenotypes. Odds ratios, 95% confidence intervals, and p-values are shown. Models were adjusted for age, sex, education years, race, ethnicity, and APOE e4 carrier status. Low cSA/low cTH was associated with the greatest odds of ADRD, exceeding odds observed for either low cSA or low cTH alone. Abbreviations: ADNI, Alzheimer’s Disease Neuroimaging Initiative; ADRD, Alzheimer’s disease and related dementias; APOE, apolipoprotein E; CI, confidence interval; cSA, cortical surface area; cTH, cortical thickness; CU, cognitively unimpaired; DMN, Default Mode Network; MCI, mild cognitive impairment; OR, odds ratio.

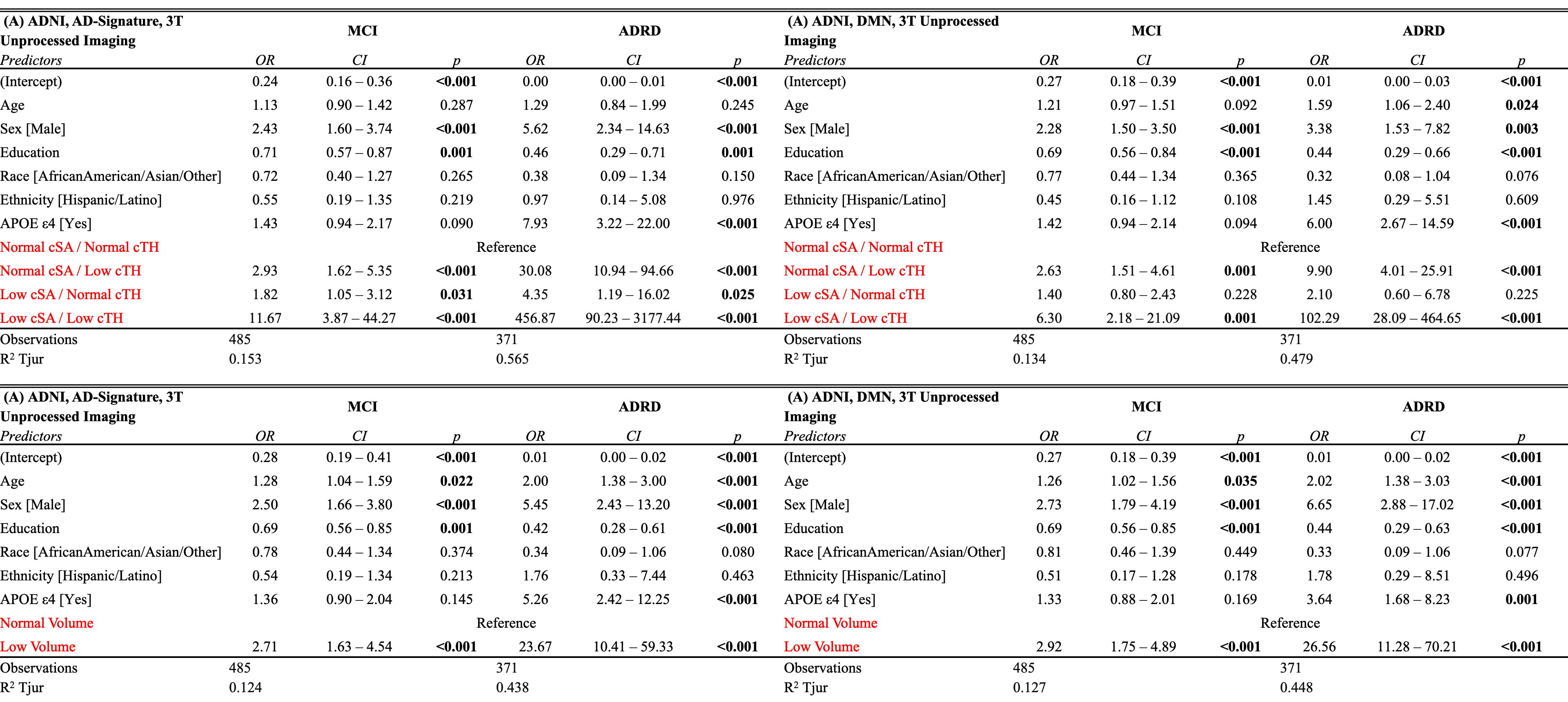

**Supplemental Figure 21. Full models for sensitivity analysis of AD-Signature and DMN surface area/thickness phenotypes and volume-defined groups in ADNI participants with 3T unprocessed imaging.** Logistic regression models show associations of cSA/cTH phenotypes and volume-defined groups with cognitive diagnosis for AD-Signature (A-B) and DMN (C-D). Models were fit separately for cSA/cTH phenotypes and volume-defined groups. CU participants served as the reference group. Odds ratios, 95% confidence intervals, and p-values are shown. Models included age, sex, education years, race, ethnicity, APOE e4 carrier status, and the morphometric phenotype or volume group. Low cSA/low cTH was associated with the greatest odds of MCI and ADRD. Low volume was also associated with greater odds of MCI and ADRD. Abbreviations: ADNI, Alzheimer’s Disease Neuroimaging Initiative; ADRD, Alzheimer’s disease and related dementias; APOE, apolipoprotein E; CI, confidence interval; cSA, cortical surface area; cTH, cortical thickness; CU, cognitively unimpaired; DMN, Default Mode Network; MCI, mild cognitive impairment; OR, odds ratio.

| **Baseline Characteristic** | **N = 1,558** |
| --- | --- |
| **Age** | 73 (7) |
| **Sex** |  |
| Female | 723 (46%) |
| Male | 835 (54%) |
| **Education** | 16.20 (2.69) |
| **Race** |  |
| White | 1,440 (92%) |
| Black/African American | 70 (4.5%) |
| Asian | 26 (1.7%) |
| American Indian/Alaskan Native | 2 (0.1%) |
| Native Hawaiian/Other Pacific Islander | 1 (<0.1%) |
| More than one Race | 19 (1.2%) |
| **Ethnicity** |  |
| Hispanic / Latino | 48 (3.1%) |
| Not Hispanic / Latino | 1,510 (97%) |
| **APOE4** | 642 (42%) |
| **Hypertension** | 733 (47%) |
| **Diabetes** | 162 (10%) |
| **Hyperlipidemia** | 862 (55%) |
| **Smoking** | 404 (26%) |
| **Cognitive Status** |  |
| CU | 659 (42%) |
| MCI | 899 (58%) |
| **Magnet Strength** |  |
| 3T | 1,014 (65%) |
| 1.5T | 544 (35%) |
| **Original/Processed Image** |  |
| Original | 319 (20%) |
| Processed | 1,239 (80%) |

**Supplemental Table 2. Baseline characteristics of the ADNI longitudinal cohort.** Baseline demographic, clinical, diagnostic, and MRI acquisition characteristics are summarized for participants included in the longitudinal analyses within ADNI participants. Continuous variables are presented as mean(SD) and categorical variables as n(%). APOE e4 carrier status was available for 1,536 participants.

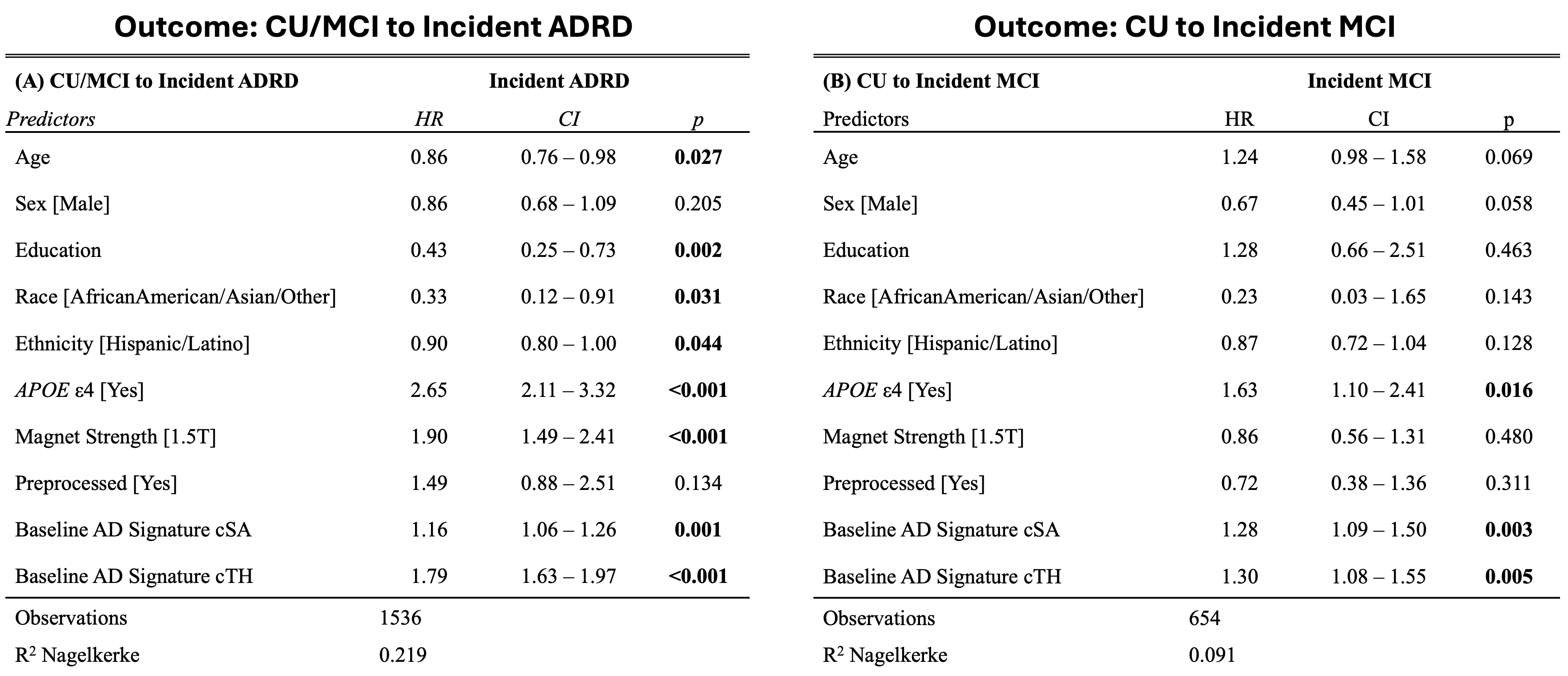

**Supplemental Figure 22. Longitudinal associations of AD-Signature surface area and thickness with incident MCI and ADRD in ADNI.** Cox proportional hazards models show associations between baseline AD-Signature cSA and cTH, and longitudinal clinical progression (A) to ADRD in baseline CU/MCI participants and (B) to MCI in baseline CU participants. Hazard ratios, 95% confidence intervals, and p-values are shown for each predictor. Models included age, sex, education years, race, ethnicity, APOE e4 carrier status, magnet strength, image preprocessing status, baseline AD-Signature cSA, and baseline AD-Signature cTH. Higher baseline cSA and cTH were independently associated with greater risk of incident ADRD and MCI. Abbreviations: ADNI, Alzheimer’s Disease Neuroimaging Initiative; ADRD, Alzheimer’s disease and related dementias; APOE, apolipoprotein E; CI, confidence interval; cSA, cortical surface area; cTH, cortical thickness; CU, cognitively unimpaired; HR, hazard ratio; MCI, mild cognitive impairment.

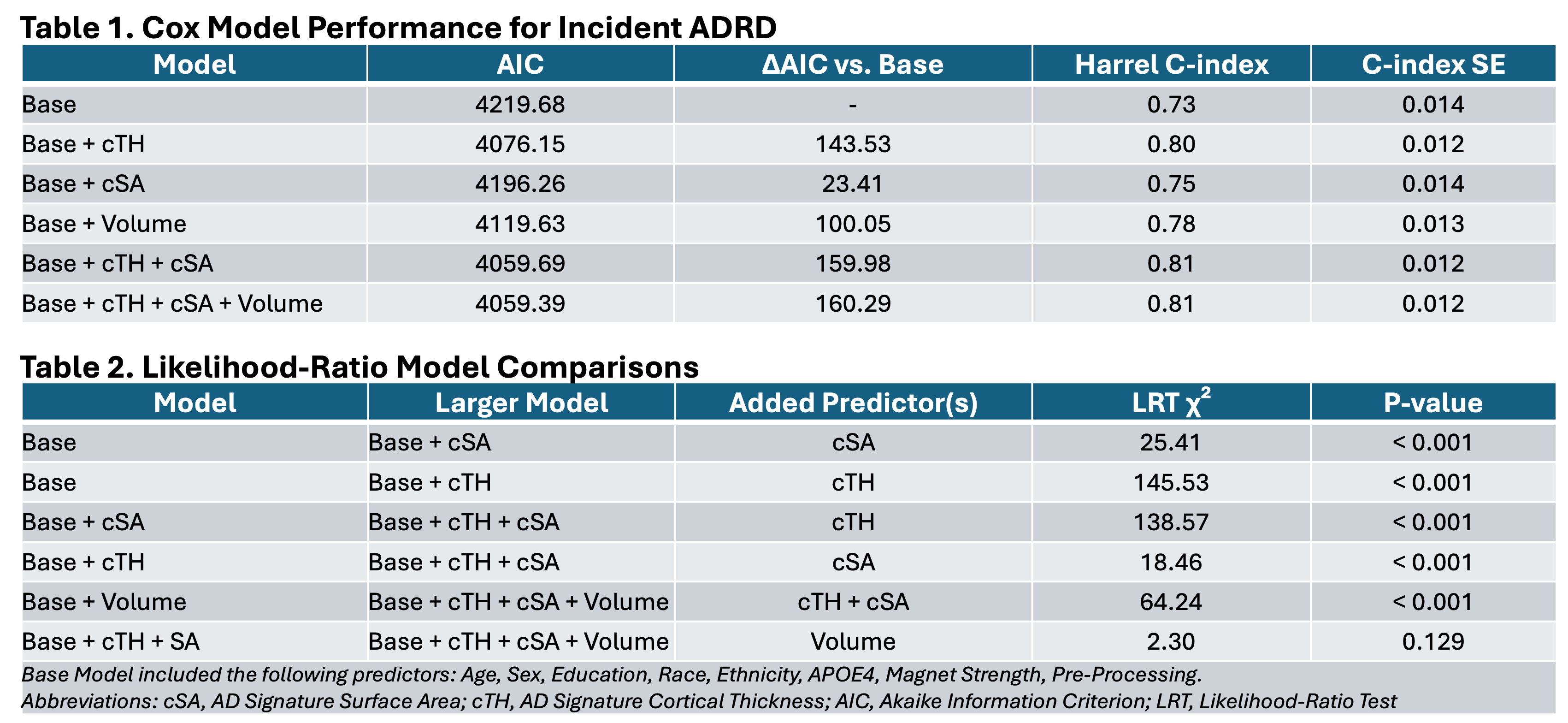

**Supplemental Figure 23. Nested Cox model performance and likelihood-ratio comparisons for incident ADRD prediction in ADNI.** Cox proportional hazards model performance was evaluated for prediction of incident ADRD among ADNI participants who were CU or MCI at baseline. The base model included age, sex, education years, race, ethnicity, APOE e4 carrier status, magnet strength, and image preprocessing status. Table 1 summarizes model fit and discrimination after adding AD-Signature cTH, cSA, volume, or combined morphometric predictors. Models including cSA and cTH had better model fit than those including volume. Table 2 shows likelihood-ratio test comparisons between nested models. Addition of cSA, cTH, or combined cSA/cTH significantly improved model fit compared to the base model. Volume did not significantly improve the cSA and cTH model, where adding cSA and cTH to the volume model significantly improved fit. Abbreviations: ADNI, Alzheimer’s Disease Neuroimaging Initiative; ADRD, Alzheimer’s disease and related dementias; AIC, Akaike information criterion; APOE, apolipoprotein E; cSA, cortical surface area; cTH, cortical thickness; CU, cognitively unimpaired; LRT, likelihood-ratio test; SE, standard error.

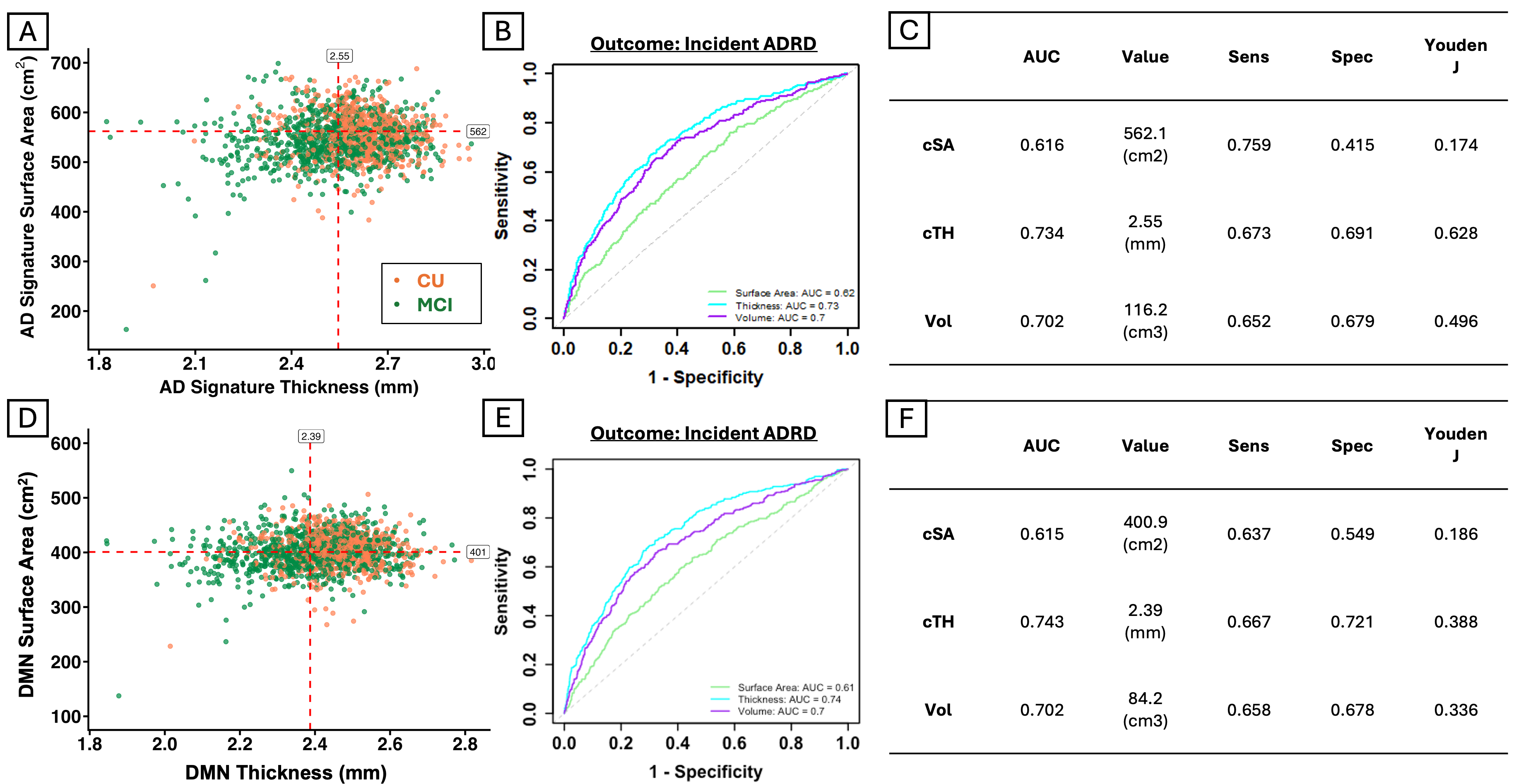

**Supplemental Figure 24. ROC-derived AD-Signature and DMN morphometric cut points for incident ADRD prediction in ADNI.** Scatterplots show baseline cSA and cTH distributions among ADNI participants who were CU or MCI at baseline for the AD-Signature (A) and DMN (D) composites. Corresponding tables summarize AUC values, optimal Youden-derived thresholds, sensitivity, specificity, and Youden J statistics for each morphometric measure for AD-Signature (B-C) and DMN (E-F) composites. Abbreviations: ADNI, Alzheimer’s Disease Neuroimaging Initiative; ADRD, Alzheimer’s disease and related dementias; AUC, area under the curve; cSA, cortical surface area; cTH, cortical thickness; CU, cognitively unimpaired; DMN, Default Mode Network; MCI, mild cognitive impairment; ROC, receiver operating characteristic.

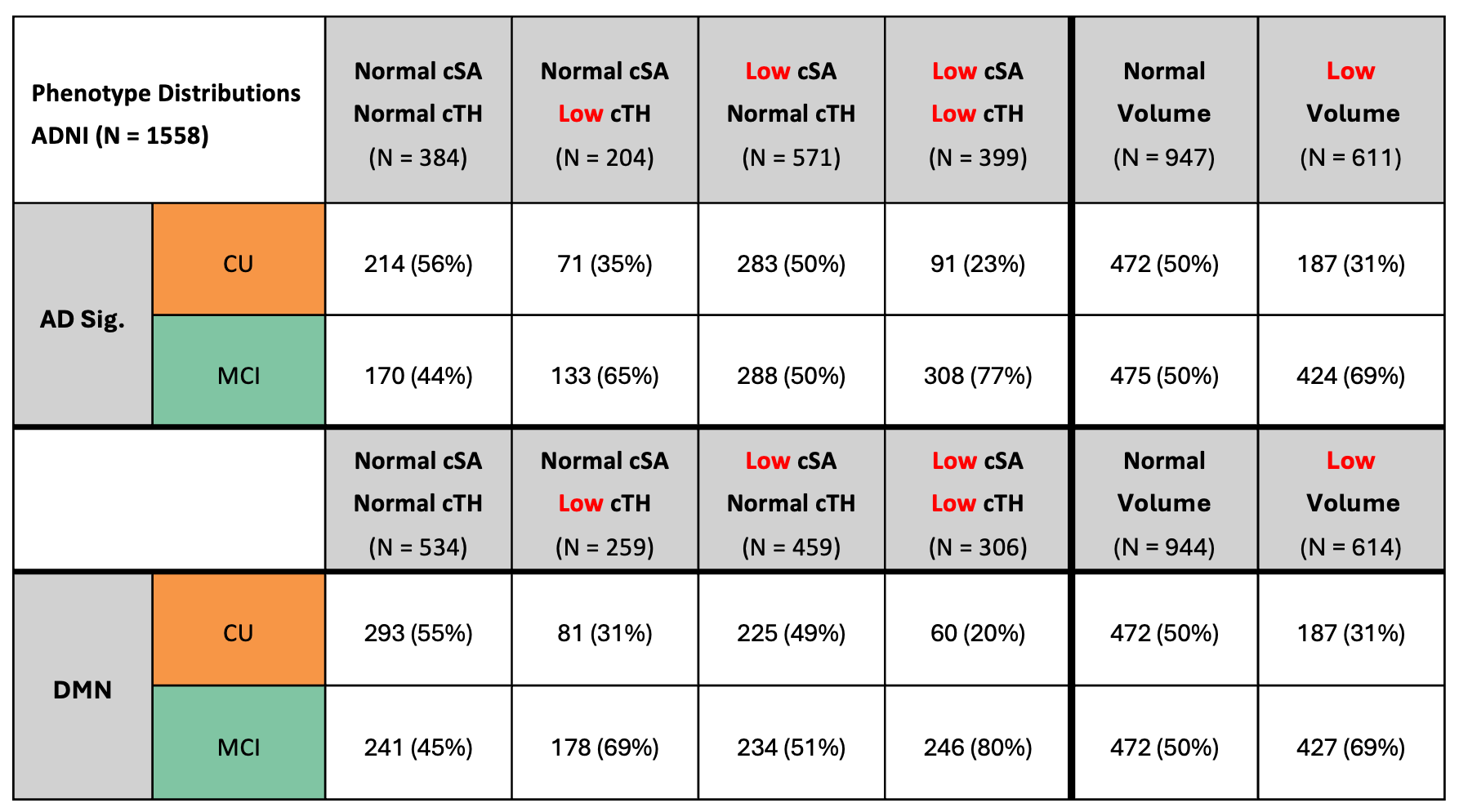

**Supplemental Figure 25. Baseline demographic distribution of ADNI longitudinal participants by morphometric phenotype.** Table shows the baseline cognitive status distribution of ADNI participants included in longitudinal analyses of incident ADRD. Participants were CU or MCI at baseline and were classified using ROC-derived cut points for cSA, cTH, and volume. Phenotype distributions are shown separately for the AD-Signature (top) and DMN (bottom) composites. Abbreviations: ADNI, Alzheimer’s Disease Neuroimaging Initiative; ADRD, Alzheimer’s disease and related dementias; cSA, cortical surface area; cTH, cortical thickness; CU, cognitively unimpaired; DMN, Default Mode Network; MCI, mild cognitive impairment.

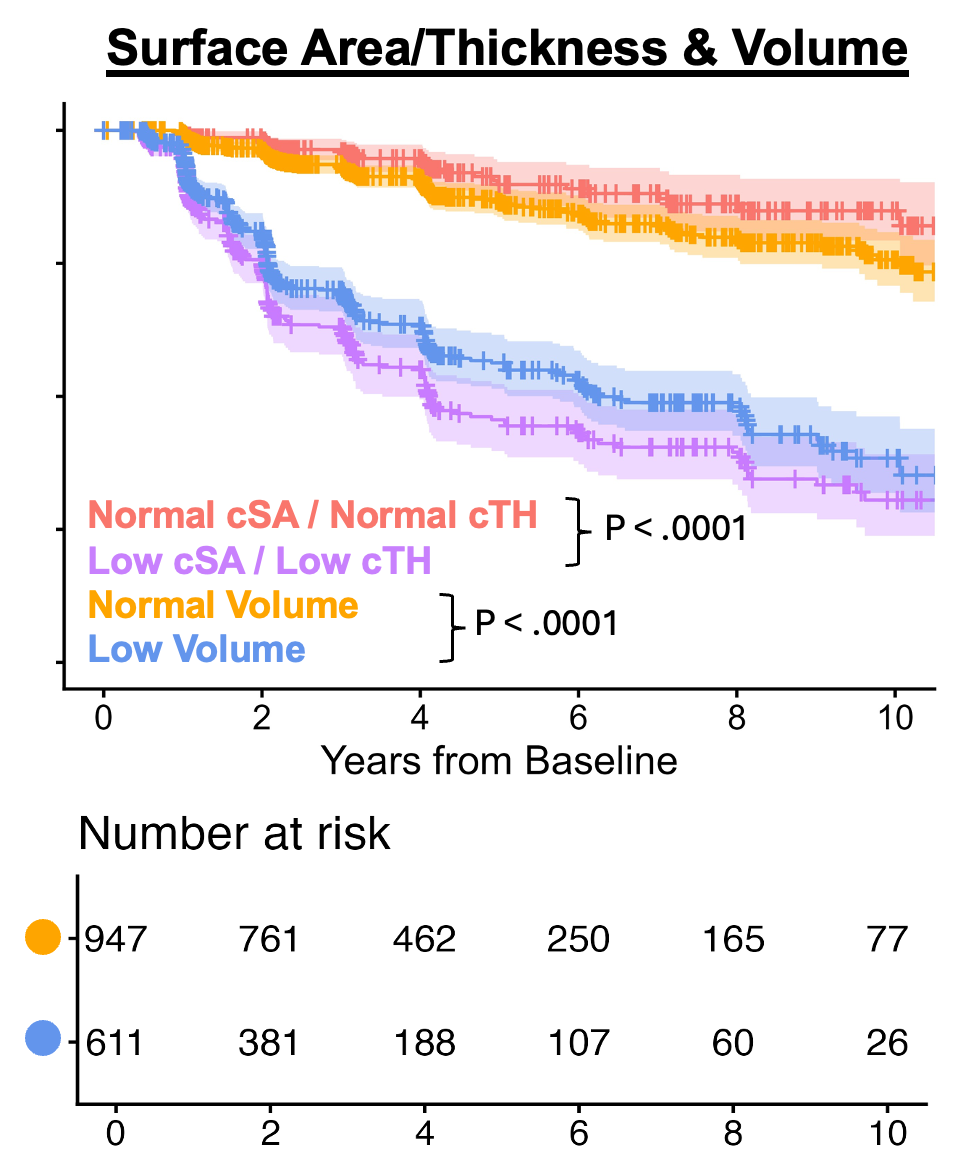

**Supplemental Figure 26. Number-at-risk table for longitudinal ADNI survival analysis of high/low volume.** Kaplan-Meier curves show ADRD-free survival among ADNI participants who were CU or MCI at baseline, stratified by AD-Signature cSA/cTH phenotype and volume group. This supplemental figure provides the number-at-risk table corresponding to Figure 4D for the high/low volume groups across 10 years of follow-up. Number at risk for cSA/cTH phenotypes can be found in Figure 4C. Log-rank p-values are shown for comparison of the high cSA/high cTH versus low cSA/low cTH phenotypes and high volume versus low volume groups. Shaded bands represent 95% confidence intervals. Abbreviations: ADNI, Alzheimer’s Disease Neuroimaging Initiative; ADRD, Alzheimer’s disease and related dementias; cSA, cortical surface area; cTH, cortical thickness; CU, cognitively unimpaired.

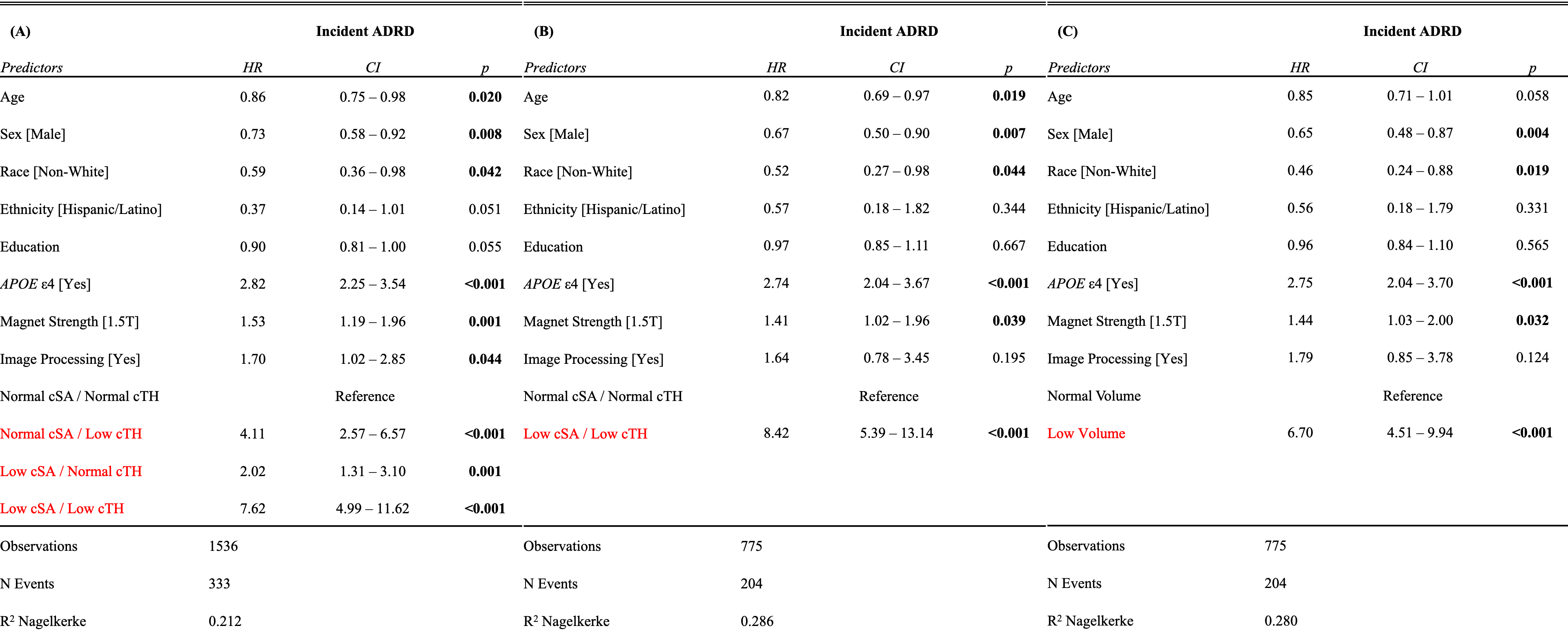

**Supplemental Figure 27. Full Cox models corresponding to Figure 4E-F.** Cox proportional hazards models show risk of incident ADRD among ADNI participants who were CU or MCI at baseline. These models correspond to the longitudinal phenotype and volume comparisons shown in Figure 4E-F. Table A includes the four-level AD-Signature cSA/cTH phenotype variable, with high cSA/high cTH serving as the reference group. Table B directly compares low cSA/low cTH to the high cSA/high cTH phenotype, excluding intermediate phenotypes. Table C compares low versus high AD-Signature volume. All models included age, sex, education years, race, ethnicity, APOE e4 carrier status, magnet strength, and image processing status. The low cSA/low cTH phenotype and low volume group were associated with greater risk of incident ADRD, with the strongest hazard observed for the combined low cSA/low cTH phenotype. Abbreviations: ADNI, Alzheimer’s Disease Neuroimaging Initiative; ADRD, Alzheimer’s disease and related dementias; APOE, apolipoprotein E; CI, confidence interval; cSA, cortical surface area; cTH, cortical thickness; CU, cognitively unimpaired; HR, hazard ratio.

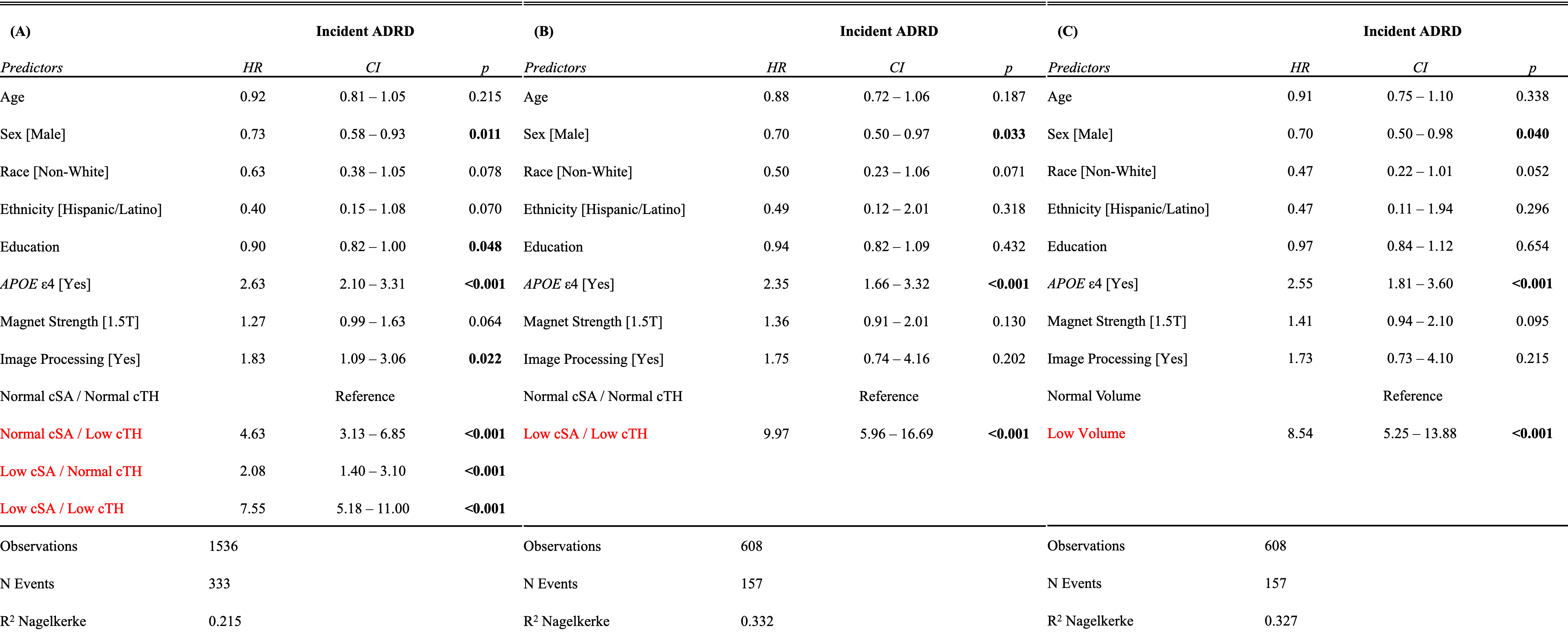

**Supplemental Figure 28. Full Cox models assessing DMN cortical surface area/thickness phenotype and volume group to incident ADRD.** Cox proportional hazards models show risk of incident ADRD among ADNI participants who were CU or MCI at baseline using DMN morphometric groups. Table A includes the four-level DMN cSA/cTH phenotype variable, with high cSA/high cTH serving as the reference group. Table B directly compares the low cSA/low cTH phenotype with the high cSA/high cTH phenotype after excluding intermediate phenotypes. Table C compares low versus high DMN volume. Hazard ratios, 95% confidence intervals, and p-values are shown for each predictor. Models were adjusted for age, sex, education years, race, ethnicity, APOE e4 carrier status, magnet strength, and image preprocessing status. The low cSA/low cTH phenotype and low volume group were associated with greater risk of incident ADRD, with the combined low cSA/low cTH phenotype showing the strongest hazard. Abbreviations: ADNI, Alzheimer’s Disease Neuroimaging Initiative; ADRD, Alzheimer’s disease and related dementias; APOE, apolipoprotein E; CI, confidence interval; cSA, cortical surface area; cTH, cortical thickness; CU, cognitively unimpaired; DMN, Default Mode Network; HR, hazard ratio.

| **Characteristic** | **N = 694** |
| --- | --- |
| **Age (years)** | 74 (7) |
| **Sex** |  |
| Female | 300 (43%) |
| Male | 394 (57%) |
| **Education (years)** | 15.89 (2.83) |
| **Race** |  |
| Asian | 9 (1.3%) |
| Black/African American | 19 (2.7%) |
| More than one Race | 3 (0.4%) |
| Native Hawaiian/Other Pacific Islander | 1 (0.1%) |
| White | 662 (95%) |
| **Ethnicity** |  |
| Hispanic / Latino | 19 (2.7%) |
| Not Hispanic / Latino | 675 (97%) |
| **APOE ε4** | 340 (49%) |
| **Hypertension** | 331 (48%) |
| **Diabetes** | 60 (8.6%) |
| **Hyperlipidemia** | 340 (49%) |
| **Smoking** | 184 (27%) |
| **Cognitive Status** |  |
| ADRD | 136 (20%) |
| MCI | 392 (56%) |
| CU | 166 (24%) |
| **Magnet Strength** |  |
| 1.5T | 241 (35%) |
| 3T | 453 (65%) |
| **Original/Processed Image** |  |
| Processed | 694 (100%) |
| **MRI-SomaScan Exam Date Difference (days)** | 89 (281) |

**Supplemental Table 3. Baseline demographics of the ADNI proteomics analysis subgroup.** Baseline demographic, clinical, diagnostic, MRI acquisition, and exam-timing characteristics are summarized for ADNI participants included in the proteomics analysis subgroup. Continuous variables are reported as mean(SD) and categorical variables as n(%). The subgroup included 694 participants with available MRI and SomaScan proteomics data. Mean difference between MRI and SomaScan exam dates was 89 days. Abbreviations: ADNI, Alzheimer’s Disease Neuroimaging Initiative; ADRD, Alzheimer’s disease and related dementias; APOE, apolipoprotein E; CU, cognitively unimpaired; MCI, mild cognitive impairment.

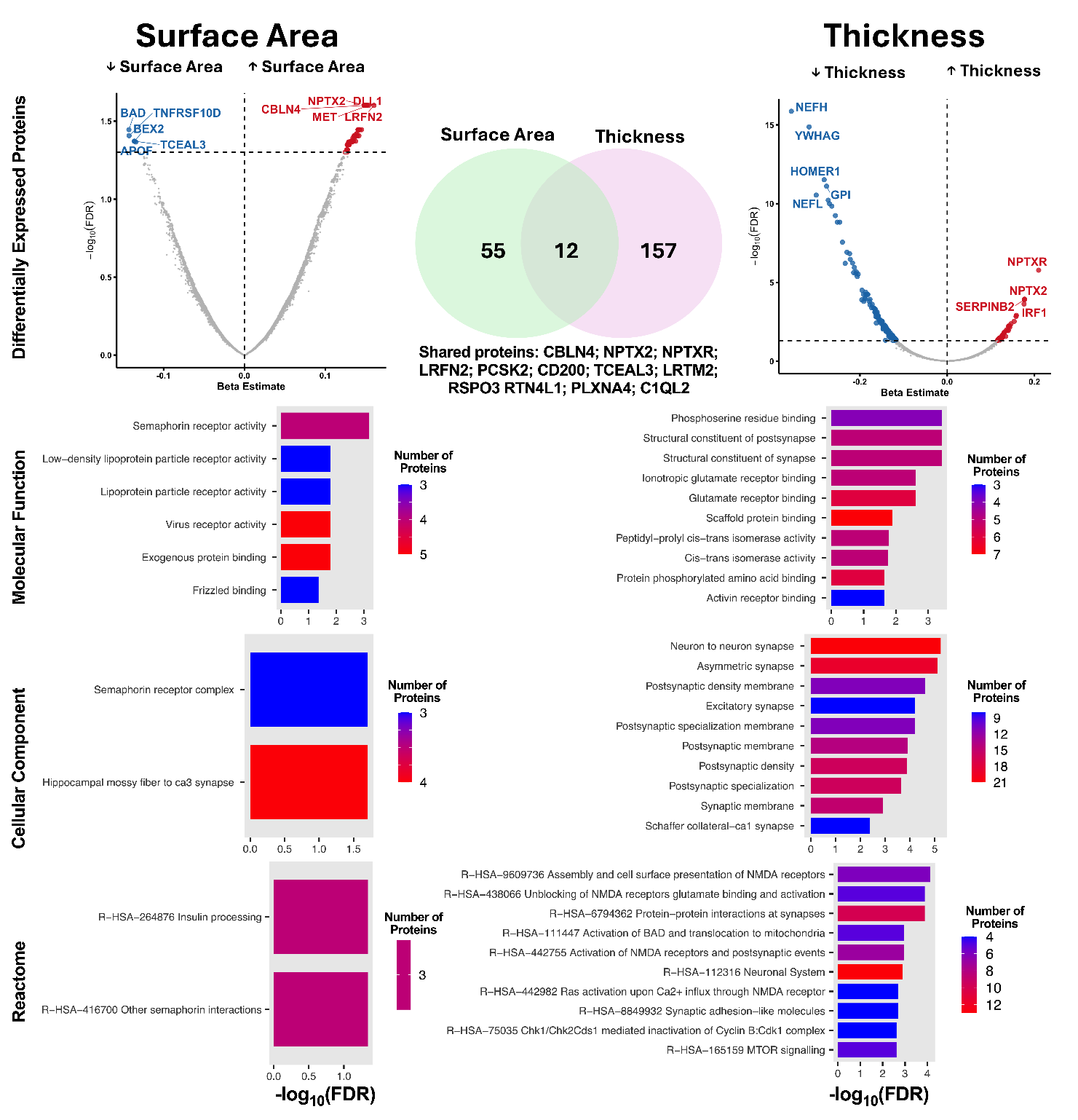

**Supplemental Figure 29. Differential proteomic signatures are observed for cortical surface area and thickness**. Multiple linear regression models adjusted for age, sex, race, ethnicity, and MRI magnet strength were utilized to identify differentially-expressed proteins (DEPs) associated with surface area and thickness, which are visualized using volcano plots. DEPs were compared between surface area and thickness using a Venn diagram. Pathway enrichment analysis was performed using DEPs for surface area and thickness as input and the Somascan 7K menu as background. The Gene ontology (GO) and Reactome datasets were queried. All DEPs and enriched pathways had an FDR *P*<.05.

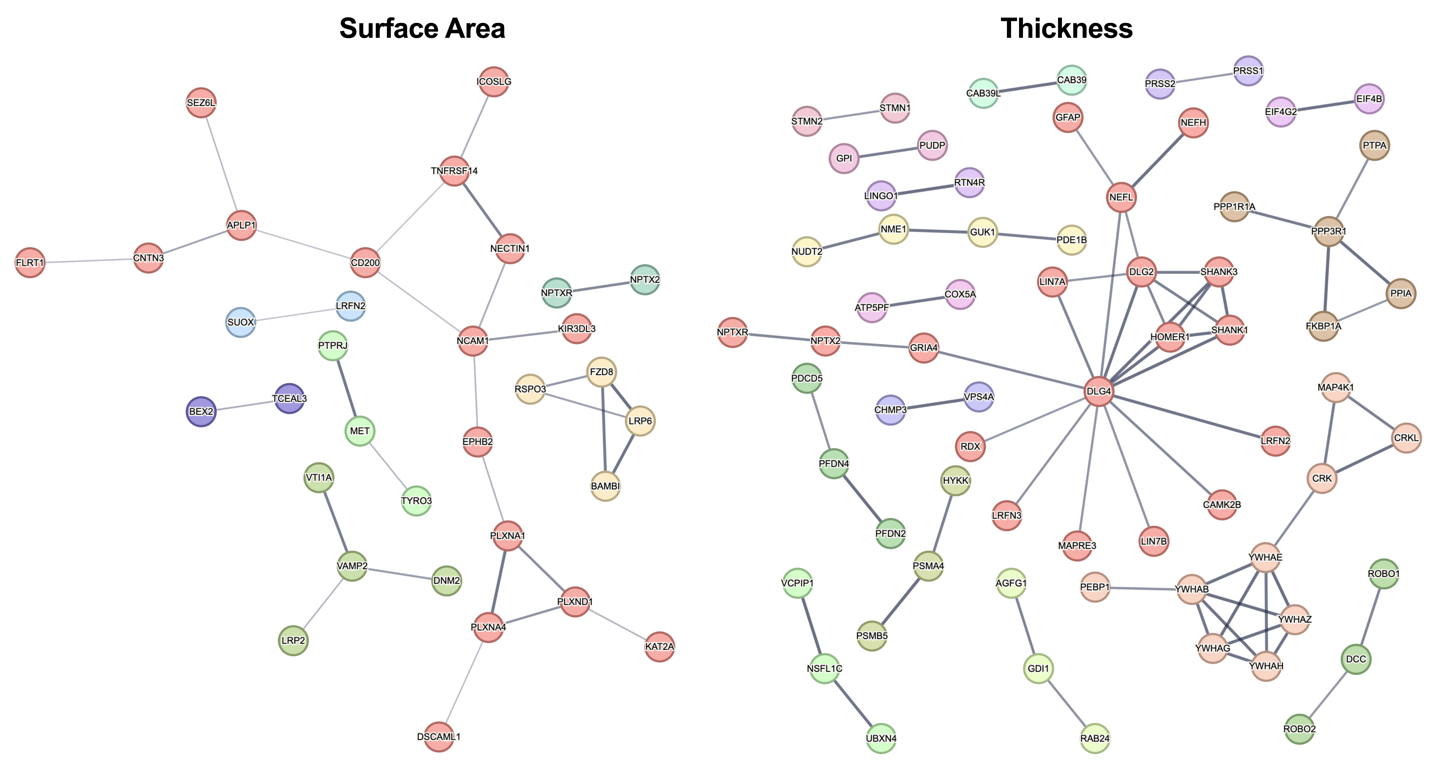

**Supplemental Figure 30. Raw protein-protein interaction networks for surface area and thickness**. Multiple linear regression models adjusted for age, sex, race, ethnicity, and MRI magnet strength were used to identify proteins associated with surface area and thickness after FDR correction, which were then visualized using protein-protein interaction networks (PPIs). PPIs had a minimum confidence of >0.4 (cSA) and >0.7 (cTH). Unconnected nodes were removed for display purposes. No first or second shell interactors were included.
